# Human GPR174 deficiency drives polyclonal lymphoproliferative disease via defects in T cell function

**DOI:** 10.64898/2026.07.14.26357774

**Authors:** Yun-Han Huang, Kathya Arana, Suzanna Rachimi, Hanson Tam, Jarmila Stremenova Spegarova, Karin R. Engelhardt, Helen Griffin, Molly Mee, Maurizio Miano, Federica Raggi, Alice Grossi, Marta Rusmini, Isabella Ceccherini, Gianluca Dell’Orso, Jacopo Ferro, Maria Carla Giarratana, Vinodh Pillai, Siddharth Banka, Tomaz Garcez, Tracy A. Briggs, Fethi Mellouli, Sandra von Hardenberg, Rita Beier, Bernd Auber, Ulrich Baumann, Hasan Tawamie, Edward Behrens, Derek A. Oldridge, Emylette Cruz Cabrera, Ying Xu, Shinyi Ouyang, Sophie Hambleton, Neil Romberg, Jason G. Cyster

## Abstract

The X-linked G-protein coupled receptor GPR174 is highly expressed in T and B lymphocytes and has immunoregulatory roles in mice, but its function in humans is unknown. We describe a cohort of six individuals who have function-disrupting variants in *GPR174* and a clinical phenotype of lymphadenopathy and autoimmunity. Histological analysis of two patient lymph nodes revealed necrotizing lymphadenitis and lymphoproliferation resembling Kikuchi-Fujimoto disease. In-depth analysis of three patients and related carriers revealed overaccumulation of CD8 terminally differentiated effector memory cells re-expressing CD45RA (T_EMRA_). Patient cells and GPR174- deficient CD8 T cells generated from controls showed less repression of proliferation by the GPR174 ligand lysophosphatidylserine (lysoPS) and an effector-biased gene expression program. GPR174-deficient CD4 T cells were resistant to lysoPS-mediated suppression of IL2 production. In mice, chronic viral infection led to over-accumulation of GPR174-deficient effector CD8 T cells. We describe an inborn error of immunity associated with dysregulated lymphocyte responses that we propose predisposes to exaggerated lymphoproliferation and autoimmunity following viral infection.

**Summary:** Six individuals with function-disrupting variants in the G-protein coupled receptor GPR174 provide insight into its immunoregulatory role. GPR174 loss emerges as a potential genetic risk factor for histiocytic necrotizing lymphadenitis (Kikuchi-Fujimoto disease) and autoimmunity via defects in T cell regulation.

## Introduction

Polyclonal lymphoproliferation is part of the normal response to infection and requires regulatory mechanisms to avoid persistent lymphadenopathy and immunopathology such as autoimmunity. Inborn errors of immunity (IEI) that genetically disrupt lymphocyte apoptosis, survival, or activation – like Autoimmune Lymphoproliferative Syndrome (ALPS), Activated PI3K Delta Syndrome (APDS), and CTLA4 haploinsufficiency – share clinical features of lymphadenopathy and autoimmunity (Costagliola et al., 2025). Causes of other lymphadenopathy syndromes remain unidentified. Histiocytic necrotizing lymphadenitis (Kikuchi-Fujimoto or Kikuchi Disease, KFD), for example, is a syndrome of cervical lymphadenopathy, fever, and associated autoimmunity. It does not have a well-defined cause, although a viral trigger has been postulated. The diagnosis relies on characteristic lymph node pathology.

Investigation of idiopathic lymphadenopathy syndromes has the potential to provide valuable insight into the mechanisms that regulate protective lymphocyte responses and defend against pathogenic responses.

*GPR174* is an X chromosome gene with a single coding exon that encodes a G-protein coupled receptor (GPCR). The gene has only one validated transcript and is primarily expressed by lymphocytes. Its gene metrics such as the probability of loss-of-function intolerance score (pLI = 0.09), loss-of-function observed/expected upper bound fraction score (LOUEF = 0.874), and missense z-score (0.91) all suggest a low probability of purifying selection against *GPR174* loss of function within the population (Karczewski et al., 2020). However, this gene may still play a role in disease pathogenesis in certain genetic and environmental contexts.

The ligand of GPR174 is lysophosphatidylserine (lysoPS), a phosphatidylserine derivative abundant in lymphoid tissues and associated with sites of inflammation and cell death (Barnes et al., 2015; Domínguez Conde et al., 2022; Heng et al., 2008; Liang et al., 2023; Liu et al., 2023). Upon receptor activation, G_s_ engages adenylyl cyclase, which increases intracellular cyclic AMP (cAMP) and leads protein kinase A to phosphorylate downstream effectors, including the transcription factor cAMP response element binding protein (CREB) (Rosenbaum et al., 2009). G_s_-coupled GPCRs and cAMP signaling tend to exert suppressive effects on lymphocyte activation and proliferation (Kammer, 1988; Raker et al., 2016).

Prior work in GPR174-deficient mice has demonstrated that GPR174 suppresses CD4 and CD8 T cell proliferation *in vitro* and in specific contexts *in vivo* and has a mild restraining effect on T_reg_ cell accumulation and function (Barnes and Cyster, 2018; Barnes et al., 2015; Shinjo et al., 2017). In B cells, GPR174 is a strong regulator of gene expression *in vitro* but the context(s) for such an effect *in vivo* remain unclear (Wolf et al., 2022). GPR174-deficient mice maintained in a specific pathogen free (SPF) environment are healthy. However, reflective of opposing effects on regulatory and inflammatory lymphocyte populations, altering the balance of GPR174 sufficient and deficient lymphocytes results in either exacerbation or attenuation of inflammatory disease modeled by experimental autoimmune encephalitis (Barnes et al., 2015; Zhao et al., 2020).

In this study, we identify a cohort of individuals with function-disrupting variants in GPR174 who have expansion of lymphoid tissues and variably penetrant autoimmunity. The clinical phenotype of the patients bearing *GPR174* loss-of-function variants suggests that GPR174 suppresses polyclonal lymphoproliferation and autoimmune risk in humans. We provide evidence through *in vitro* analysis of GPR174-deficient human T cells and *in vivo* assessment of the anti-viral CD8 T cell response in GPR174-deficient mice, that T cell dysfunction contributes to disease pathogenesis.

## Results

### Identification of individuals with immune-mediated disease and GPR174 variants

During the COVID-19 pandemic, a boy (P3) between the ages of 1 and 5 developed a particularly severe case of COVID-19 complicated by *Staphylococcus aureus* lymphadenitis, autoimmune hemolytic anemia (AIHA), and severe idiopathic thrombocytopenic purpura (ITP). Despite intravenous immunoglobulin (IVIG), high dose steroids, daily platelet infusions, and eltrombopag (thrombopoietin receptor agonist) treatment, he died of intracerebral hemorrhage while awaiting hematopoietic stem cell transplantation. Because COVID-19 infections in children are usually mild but severe infections can develop in those with inborn errors of immunity (Zhang et al., 2020), we performed trio whole genome sequencing, identifying a maternally inherited nonsense variant, p.R53X (NM_032553, c.157C>T), in *GPR174*. No additional significant variants were identified in genes associated with Fanconi anemia, leukemia, or known IEI (International Union of Immunological Societies Inborn Errors of Immunity gene list) (Poli et al., 2025).

Facilitated by GeneMatcher (Sobreira et al., 2015), we ascertained five more individuals from four families in Europe and the United States with rare, nonsynonymous, hemizygous variants in *GPR174* and immune-mediated disease (Fig 1A, B). There are four unique variants amongst the six individuals (P1-6). Two variants, p.R18X (c.52C>T) and p.R53X, have truncations within the first two of the seven transmembrane domains. Two variants are missense mutations, p.R75K (c.224G>A) and p.R302W (c.904C>T). Two unrelated individuals have the p.R18X variant and two brothers share the p.R75K variant. All the variants are predicted to be structurally damaging. The Combined Annotation Dependent Depletion (CADD) scores of the variants are above 20, placing them within the top 1% of predicted deleterious variants (Schubach et al., 2024), and the minor allele frequencies are <0.0001 (ultra-rare) (Akalu and Bogunovic, 2024) or undetected within the public databases that we queried (Fig 1B). None of the individuals have other predicted deleterious variants in known immune disease associated genes. Together, these observations suggest a high likelihood that the *GPR174* variants are disease-causing.

**Figure 1:**
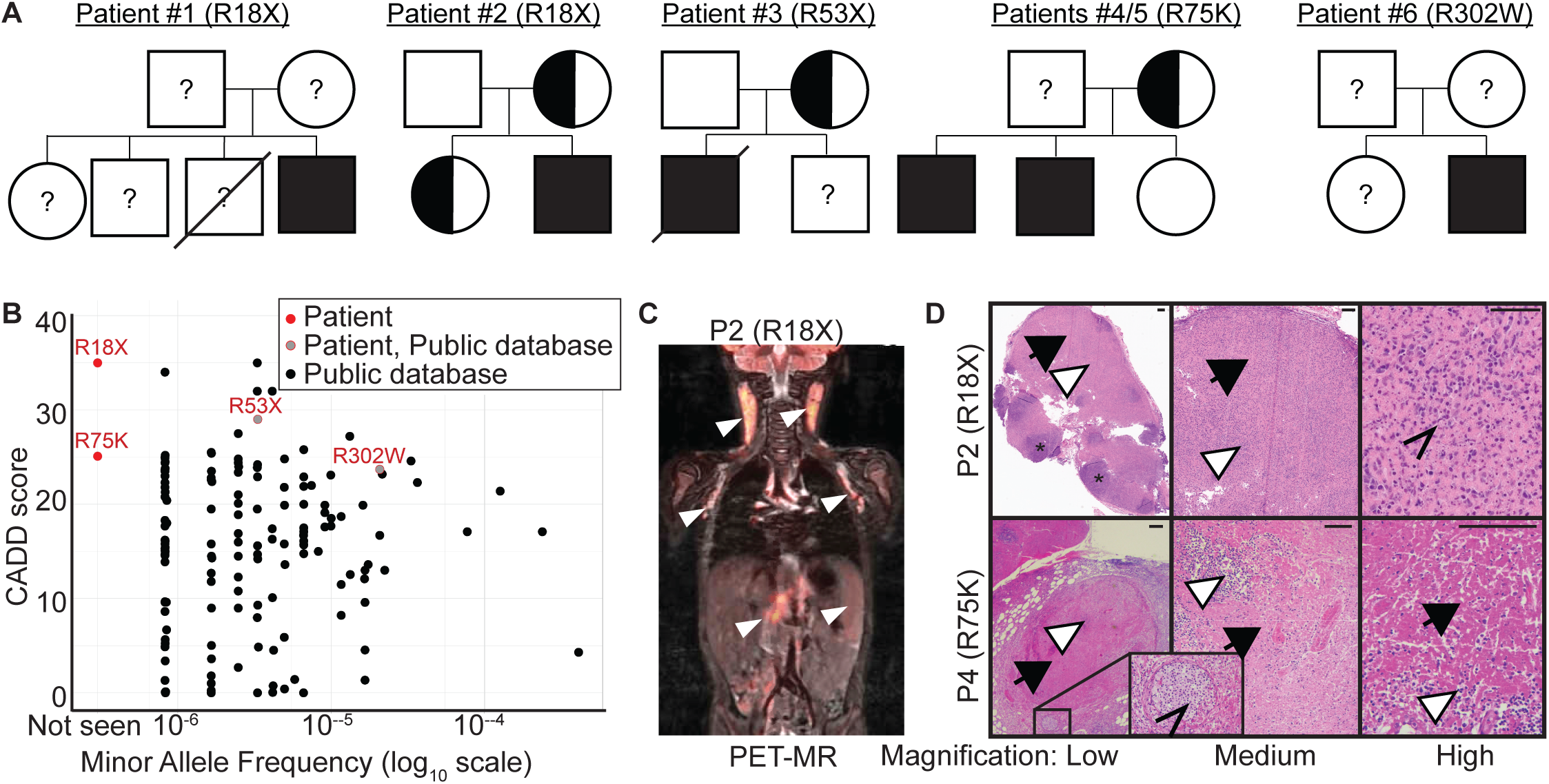
Description of GPR174-deficient patients. A: Pedigrees of the patient cohort. B: CADD vs gnomAD allele frequencies. Red labels indicate variants of patients within our cohort. C: PET-MR scan of P2. White arrowheads denote areas of hypermetabolic uptake. D: Hematoxylin & eosin stain of cervical lymph node biopsies of P2 and P4. Black arrowheads indicate areas of necrosis. White arrowheads indicate lymphocytic infiltrate. Carets indicate histiocytes. Asterisks indicate B cell follicles. Scale bar = 100 µm.

Phenotypically, all the individuals have had clinically significant chronic or recurrent lymphoid hyperplasia in the form of lymphadenopathy, tonsillar expansion, and/or splenomegaly (Table 1, Fig 1C). Lymph node biopsies were performed in five of the six individuals and did not identify malignancy. Other than *Staphylococcus aureus* lymphadenitis diagnosed in P3, no other infection was identified. Histologic examination of cervical lymph node biopsies from P2 and P4 demonstrated necrotizing lymphadenitis with lymphoid infiltration, associated histiocytes, and absent neutrophils, consistent with histiocytic necrotizing lymphadenitis (Fig 1D). A second retroperitoneal lymph node biopsy of P4 three years after the cervical lymph node biopsy demonstrated consistent pathology. Repeat cervical lymph node biopsy from P2 one year after the initial biopsy demonstrated interfollicular lymphoid expansion (Supp Fig 1A), consistent with a proliferative phenotype hypothesized to be a precursor phase of histiocytic necrotizing lymphadenitis. Together, the lymph node biopsies implicate an inflammatory process resembling that of KFD.

**Table 1:**
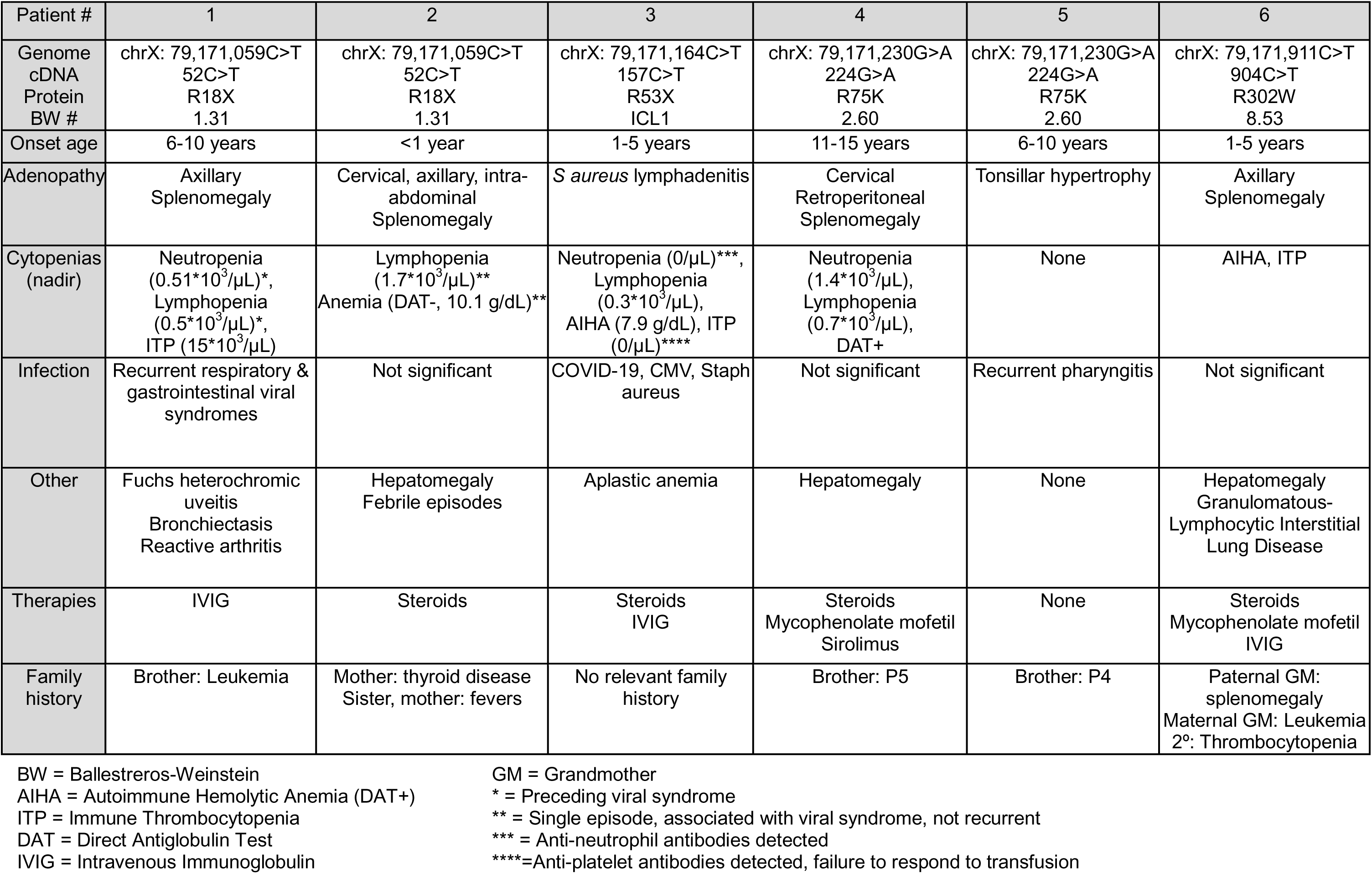
Clinical characteristics of GPR174 deficient patients.

In addition to lymphoid hyperplasia, three patients have had unusually frequent or severe infections, three have experienced autoimmune cytopenias, and four have had other organ-specific inflammation, particularly in the lungs and liver (Table 1, Supp Fig 1B). Inflammatory episodes have been successfully treated with immunosuppressive agents including steroids, IVIG, mycophenolate mofetil, and sirolimus (see Supp Table 1 for case details). While autoimmune lymphoproliferative syndrome (ALPS) was considered within the differential diagnosis of these patients, and double-negative TCRαβ ^+^ T cells were elevated in three individuals (P2, P4 and transiently P5; Table 2), no germline pathogenic variants within known ALPS-related genes were detected and a lymphocyte apoptosis assay was negative in P4. P1 has an immunophenotype consistent with common variable immunodeficiency (CVID) with low immunoglobulins, defective vaccine responses, low class-switched memory B cells, and clinically appropriate infections; genetic variants in known CVID-associated genes were not detected. The conjunction of these clinical and genetic findings in multiple unrelated individuals strongly suggested that immune dysregulation with features of autoimmunity and altered host defense was causally associated with the *GPR174* variants.

**Table 2:**
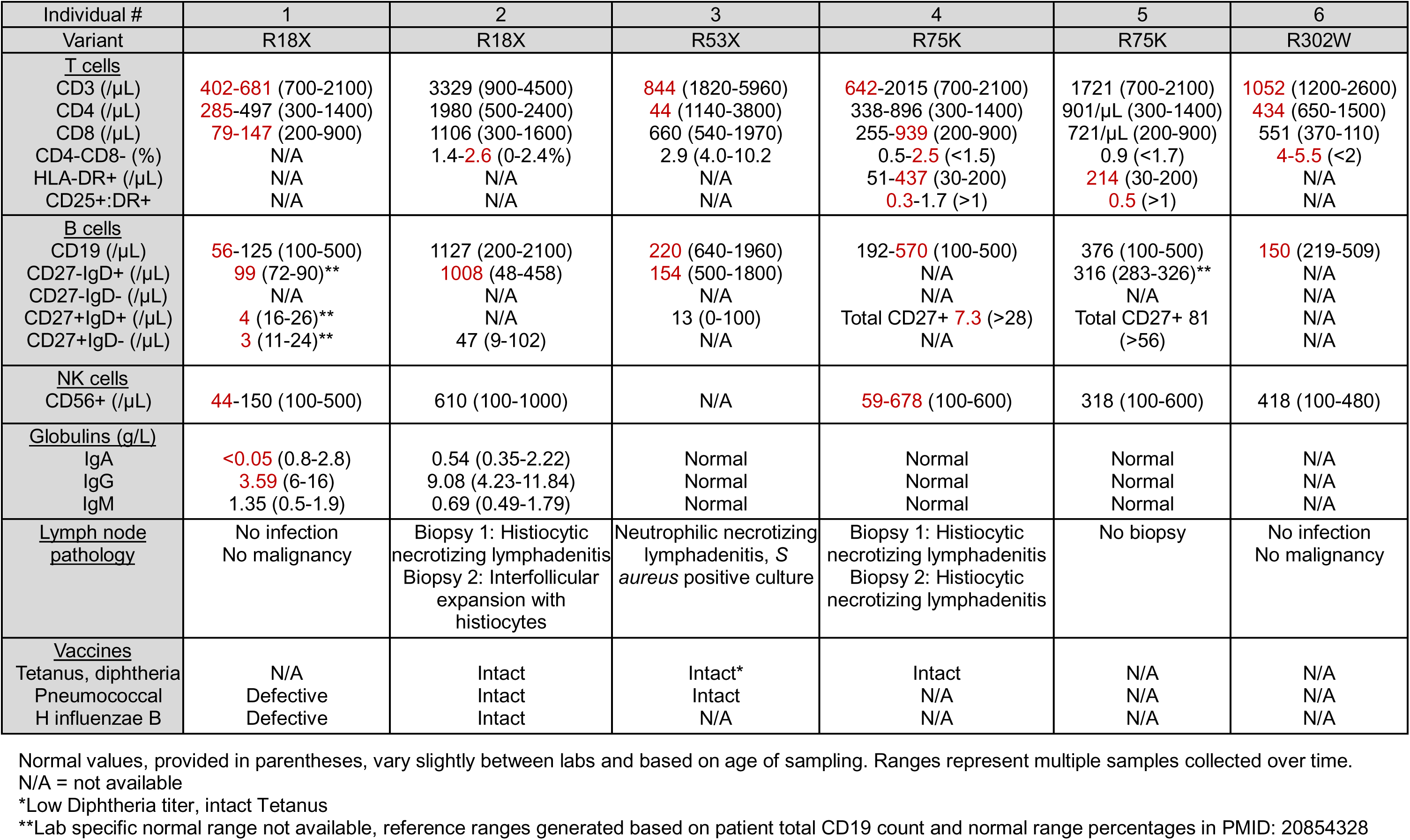
Clinical immunophenotypes of GPR174 deficient patients.

### GPR174 disease-associated variants are loss-of-function

The amino acid disruptions of the GPR174 variants suggest that they are most likely loss of function (Fig 1B, 2A). When modelled into the cryo-EM structure of GPR174 (Liang et al., 2023), the two truncating variants (R18X and R53X) lie within the first two transmembrane domains of the seven-transmembrane receptor. R75K involves a residue previously characterized to mediate interaction with the lysoPS ligand, and the replacement of arginine’s guanidino group with an extra carbon and amine would displace the nitrogen that forms a salt bridge with lysoPS (Liang et al., 2023; Liu et al., 2023; Nie et al., 2023). R302W is at the junction between the final transmembrane domain and the C-terminal regulatory region and alters a positively charged hydrophilic to a large, hydrophobic residue. Like the truncations, the patient-derived missense variants therefore may interrupt important functional elements of GPR174.

To directly assess the functional consequences of each of the GPR174 variants, we introduced wild-type (WT) and variant receptors into a 293T cell line with a cAMP-responsive element reporter (Fig 2B, Supp Fig 2A). This previously described system (Howard et al., 2025) leverages landing pad technology for single-copy insertion of the reporter and places the receptor under a doxycycline-inducible promoter. Receptor expression was titrated to minimize possible overexpression artifacts. Using this system, we observed decreased GPR174 abundance with the variants compared to the WT in total protein detected by Western blot for an N-terminal epitope (OX56) tag. There was complete loss for the truncation variants and markedly reduced protein levels for R75K and R267I (an additional variant noted in a public database whose clinical phenotype is complicated by concomitant cartilage hair hypoplasia) (Fig 2C). Consistent with the more flexible structure of the C-terminus, the R302W variant had a partial effect on protein abundance (Fig 2C). GPR174 has two predicted extracellular N-glycosylation sites (N4 and N164; Uniprot Q9BXC1) (Apweiler et al., 2004) and the variable electrophoretic mobility of the protein above the predicted 38kD molecular weight likely reflects different glycoforms or possibly receptor aggregates. The impact of the variants on receptor abundance at the cell surface as detected by flow cytometry was in close alignment with the Western blot analysis (Fig 2D). These results suggest that the missense variants, in addition to regulatory effects, disrupt receptor structure and stability.

**Figure 2:**
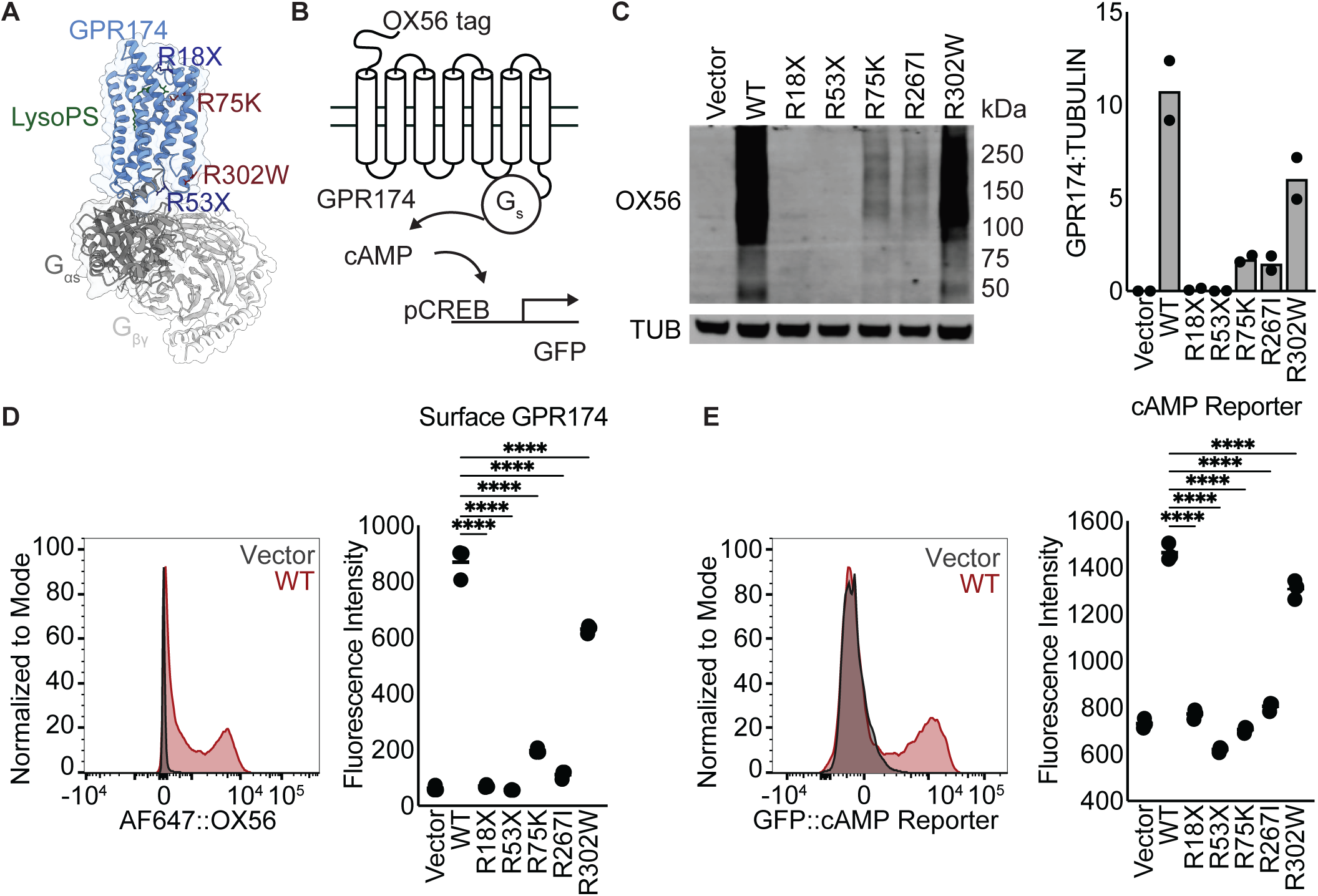
Functional characterization of patient-derived GPR174 variants. A: Ribbon diagram of lysoPS-bound GPR174 based on CryoEM structure (pdb7xv3) with variants highlighted. The associated G_s_ protein complex is shown in gray. B: Diagram of CRE reporter system with N-terminal OX56 tag and GFP transcriptional reporter. C: Total protein assessed by immunoprecipitation of OX56-GPR174 followed by Western blot detection of OX56 epitope (TUB = tubulin). Representative of two independent experiments. Graph shows summary densitometry data from two independent experiments. D: Surface protein detected by flow cytometry analysis of OX56-GPR174. Histogram plot shows example staining of Vector and WT OX56-GPR174 expressing cells. The histogram and graph of mean fluorescence intensity data are representative of three independent experiments. One-way ANOVA with Dunnett’s correction, ****=p<0.0001. E: Transcriptional activity downstream of GPR174 assessed by flow cytometric detection of a CRE-GFP reporter. Histogram plot shows example of Vector and WT OX56-GPR174 expressing cells. The histogram and graph of mean fluorescence intensity data are representative of three independent experiments. One-way ANOVA with Dunnett’s correction, ****=p<0.0001.

Receptor signaling through G_s_ via cAMP/pCREB as detected by the GFP cAMP responsive element (CRE) reporter correlated with receptor abundance. Signaling was blocked or strongly attenuated by all the variants except R302W for which there was a ∼25% reduction (Fig 2E). No added ligand was needed to observe these effects, consistent with prior studies suggesting that lysoPS present in cell culture is sufficient for receptor activation (Nie et al., 2023; Wolf et al., 2022). Together, these observations establish that all the patient-derived GPR174 variants result in decreased receptor availability and downstream signaling.

### Hemi- and heterozygous GPR174 loss favors expansion of terminally differentiated peripheral CD8 cells

GPR174 is expressed broadly across multiple subsets of lymphocytes in humans (Fig 3A) (Domínguez Conde et al., 2022; Xu et al., 2023). Notably, expression is highest in antigen-experienced CD8 T lymphocytes, particularly in tissues and lymph nodes (Fig 3A). CD4 T cells, natural killer (NK) cells, and B cells also express detectable levels of GPR174, whereas myeloid cells and plasma cells have very low levels. We hypothesized that cells highly expressing GPR174 would be most affected by GPR174 loss. Clinical flow cytometry enumerating patient lymphocyte subsets demonstrated low CD4 T cell concentrations in three patients, memory B cell deficiency in three patients, and elevated double-negative T cell frequencies in three patients (Table 2). Resolution of some of these differences upon clinical resampling suggested secondary effects from transient inflammation or temporary anti-inflammatory therapy.

**Figure 3:**
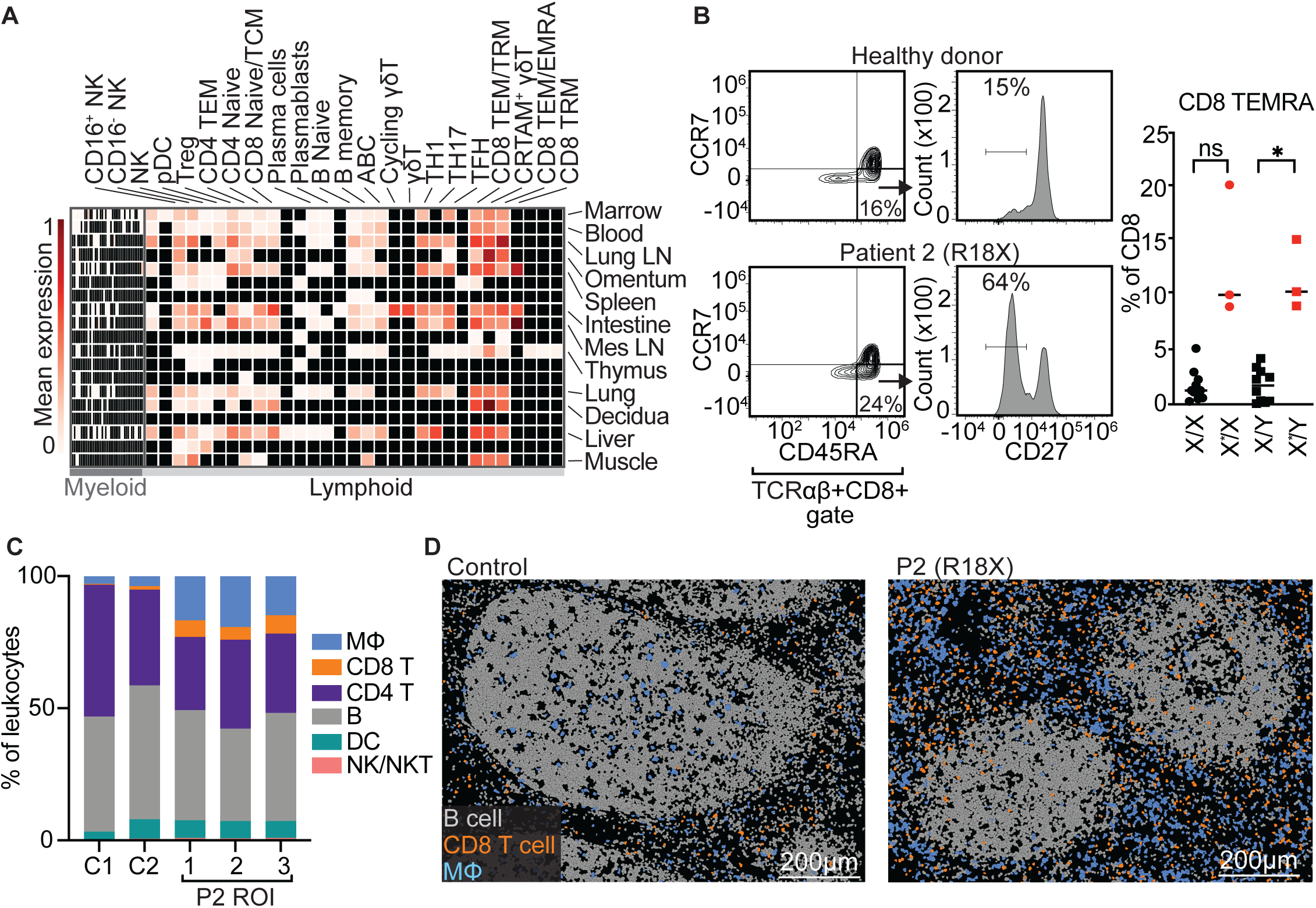
Characterization of GPR174-deficient patient blood and lymph nodes. A: Expression of *GPR174* in human cells across tissues in public single-cell datasets (Domínguez Conde et al., 2022; Xu et al., 2023). Selected cell types shown from heatmap generated at celltypist.org. Tissues and cell types with the highest expression are labeled. Black boxes indicate unavailable data. LN = Lymph Node, Mes = Mesenteric. B: Flow cytometric identification of TCRαβ^+^CD8^+^CCR7^-^CD45RA^+^CD27^-^ T_EMRA_ cells and their frequencies among total TCRαβ^+^CD8^+^ T cells in blood samples from male GPR174-deficienct patients (X*/Y; n=3), female carrier relatives (X*/X; n=3) and GPR174-sufficient male/female age-matched controls (X/Y; n=10. X/X; n=10). *p<0.05 by t-tests with Welch’s correction. C: Composition of immune cells within two control ROIs (1 ROI per donor) and P2 ROIs (3 ROIs across two lymph nodes) assessed by Xenium spatial transcriptomics. D: Positions of B cells, CD8 T cells, and macrophages in cervical lymph node biopsy #2 from P2 and a control reactive cervical lymph node from a pediatric male.

To identify the primary effects of GPR174 deficiency, we deeply immunophenotyped the peripheral blood of three affected males (P2, P4, P5) during periods of relative wellness and compared results to samples from healthy female carrier relatives (P2’s sister and mother, and P4/5’s mother) and age/sex matched healthy donors. Significant differences were not noted in lineages with minimal GPR174 expression including monocytes and dendritic cells (DCs) (Supp Fig 3A). Among lymphocytes, which express more GPR174, patients had low CD4:CD8 ratios, but no consistent frequency differences for most subsets including CD4 and CD8 naïve (T_naïve_), central memory (T_CM_), and effector memory (T_EM_) subsets as well as T regulatory (T_reg_) cells, B cell subsets, and NK cells. Most notable was a four-fold increase in the abundance of CD8+CCR7-CD45RA+CD27- T effector memory re-expressing CD45RA (T_EMRA_) cells for patients and female carrier relatives compared to age-matched controls (Fig 3B).

These findings support that GPR174 deficiency permits CD8 T_EMRA_ accumulation in both patients and carriers and suggest that additional mechanisms contribute to the inflammatory phenotype of the patients compared to the carriers.

Since *GPR174* is an X-linked gene, we queried the presence of skewed X-inactivation in the GPR174 variant carriers, which could imply a selective advantage for one allele over the other. Specifically, we hypothesized that variant GPR174-expressing cells may be enriched within the CD8 T_EMRA_ compartment and contribute to its relative expansion in clinically unaffected carriers. We therefore performed single-cell RNA sequencing (scRNAseq) of peripheral blood mononuclear cells (PBMCs) from P2’s mother and sister. We found that CD4 and CD8 T cells inactivate *GPR174* to a similar degree to well-annotated X-inactivated genes with a small increase in biallelic expression in CD4 T cells compared to CD8 T cells (Supp Fig 3B). Since our dataset did not capture sufficient B and NK cells to perform this analysis, we also queried publicly available scRNAseq datasets of healthy individuals (Domínguez Conde et al., 2022; Ke et al., 2025; Xu et al., 2023) and found a similarly strong inactivation in B and NK cells (Supp Fig 3B). While *GPR174* is annotated as a variably inactivated gene in chromosome-wide analyses of X inactivation (Balaton et al., 2015; Tomofuji et al., 2024b), our focused analyses suggest that, at homeostasis, it is subject to inactivation to a similar extent as well-annotated inactivated genes. This observation enabled us to query the CD8 T_EMRA_ population for allelic skewing. Unexpectedly, we found that this population was not skewed towards the variant allele and in fact favored skewing towards the WT allele (Supp Fig 3C, Supp Table 2). We speculate that this may be due to survival or recirculation biases in the variant versus wild-type CD8 T_EMRA_ cells, which favor detection of wild-type cells in circulation when both populations are present.

### Abundant CD8 T cells and macrophages are features of a GPR174-deficient patient’s lymph nodes

All identified GPR174-deficient patients experienced secondary lymphoid hyperplasia, so we explored how GPR174 loss affects lymph nodes. We more deeply evaluated two excisional cervical lymph node biopsies from P2. Clinical immunohistochemical (IHC) stains of biopsy #1, which was collected at age 1-5, showed areas of necrosis engorged with Ki67+ proliferating CD8 T cells and histocytes (Supp Fig 4A). Biopsy #2, collected a year later, was not necrotic, but in accord with the hematoxylin and eosin analysis (Supp Fig 1A), featured enlarged interfollicular zones containing numerous CD8 T cells and histocytes (Supp Fig 4A).

**Figure 4:**
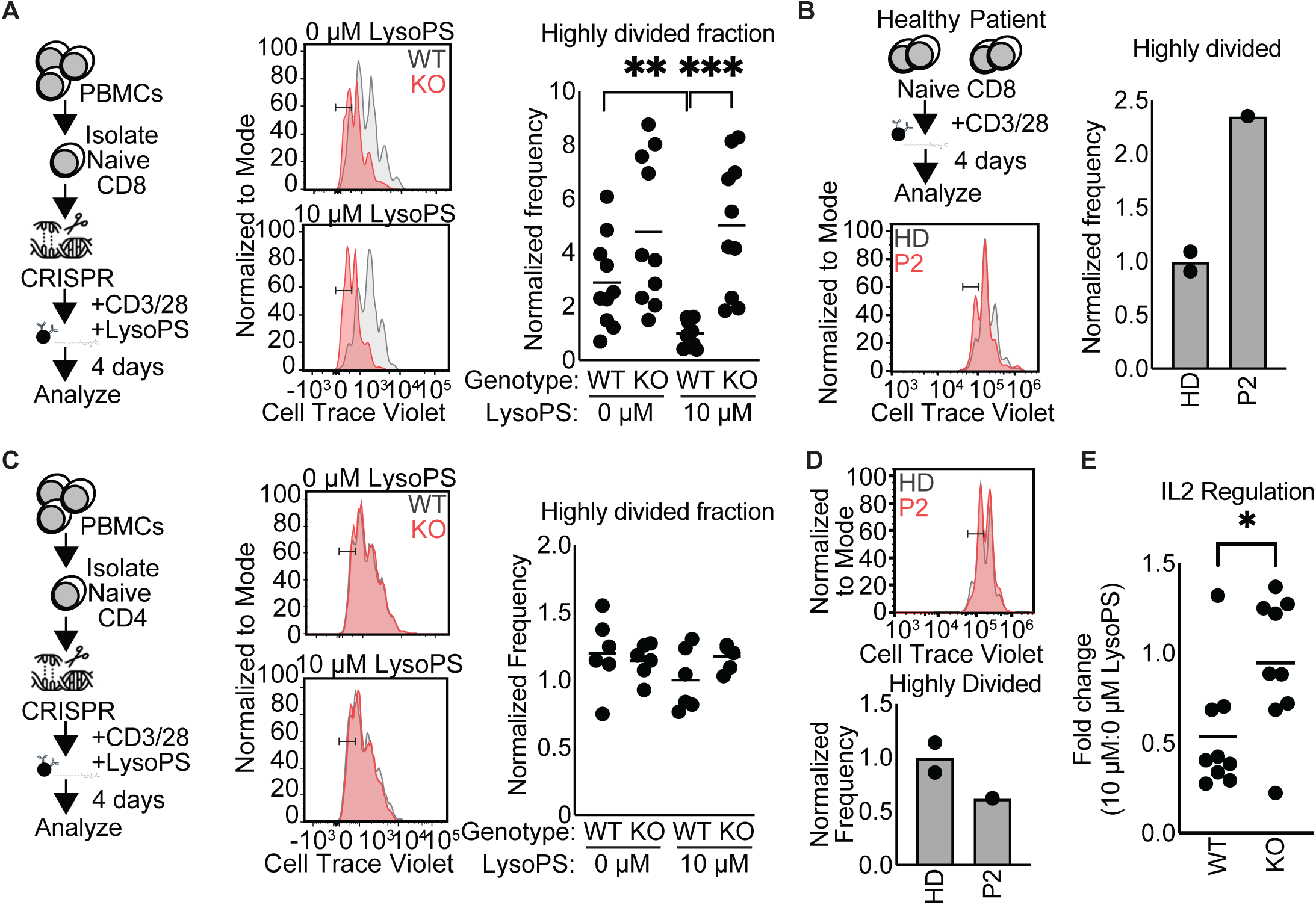
Functional analysis of GPR174-deficient lymphocytes. A: Left - Experimental scheme: CD8 T cells were edited using sgRNAs targeting *GPR174*, then stimulated with CD3/CD28 beads for 4 days +/- 10 µM lysoPS. Center – Example histogram plots of cell trace violet (CTV) dilution by stimulated CD8 T cells. Right – Summary graph showing data pooled from 5 independent experiments with 4 different donors. WT = unedited and safe-harbor targeted, KO = GPR174 sg1 and sg2. Group means are indicated. t-test with Welch’s correction, **p<0.01, ***p<0.001. B: Naïve CD8 T cells collected from healthy donors (HD) or a patient carrying the R18X variant (P2) were stimulated with CD3/CD28 beads for 4 days. Histogram plot shows example CTV dilution and graph shows normalized frequency of highly divided cells. C: CD4 T cells were edited using sgRNAs targeting GPR174, then stimulated with CD3/CD28 beads. Pooled from 3 independent experiments with 3 different donors. D: Flow cytometry analysis of naïve CD4 cells collected from a GPR174-deficient patient and stimulated with CD3/CD28 beads for 4 days compared to healthy controls. E: IL2 ELISA of the supernatant collected from the CD4 proliferation assay in C after 4 days, t-test with Welch’s correction, *p<0.05.

To resolve more cell types, their relative positions, and their biologic states, we performed spatial transcriptomic analyses on three, non-contiguous regions of interest (ROIs) from two separate lymph nodes within the P2 biopsy #2 (Supp Fig 4B).

Representative ROIs in reactive cervical lymph nodes from two pediatric GPR174- sufficient male donors served as controls. Cells within ROIs were segmented, clustered by gene expression (Supp Fig 4C), and annotated using canonical lineage-specific transcripts. CD8 T cell, CD4 T cell, B cell, macrophage, DC and NK/NKT subsets were identified for downstream compositional, positional, and transcriptional analyses (Supp Fig 4D). Consistent with the IHC staining, P2 CD8 T cells were 7-fold and macrophages 5-fold more abundant than counterpart cells in control ROIs (Fig 3C, Supp Table 3).

Two-dimensional positional analyses of these two cell types and B cells (to mark follicles) revealed numerous macrophages in P2’s lymph node, concentrated within interfollicular zones (Fig 3D). P2 CD8 T cells were also numerous and localized both within and outside of B cell follicles. In comparison, macrophages from control ROIs primarily resided at the follicular border or within follicles, whereas CD8 T cells almost exclusively localized outside of them. These findings suggest a role for dysregulation of CD8 T cells and macrophages in GPR174 deficiency disease pathogenesis, concordant with the expected immunopathology of KFD.

### GPR174 deficiency results in CD8 hyperproliferation and loss of CD4 IL2 suppression

Prior studies have demonstrated regulatory roles of GPR174 on mouse CD4, CD8, and B lymphocytes. With a particular interest in CD8 T cells given the findings in the patient samples, we assessed human T and B cell regulation with paired studies of CRISPR-edited and patient-derived lymphocytes. To functionally interrogate the role of GPR174, isolated from the effect of other genetic and epigenetic differences, we used CRISPR to knock out *GPR174* in naïve CD8 T cells from healthy donors (Supp Fig 5A).

**Figure 5:**
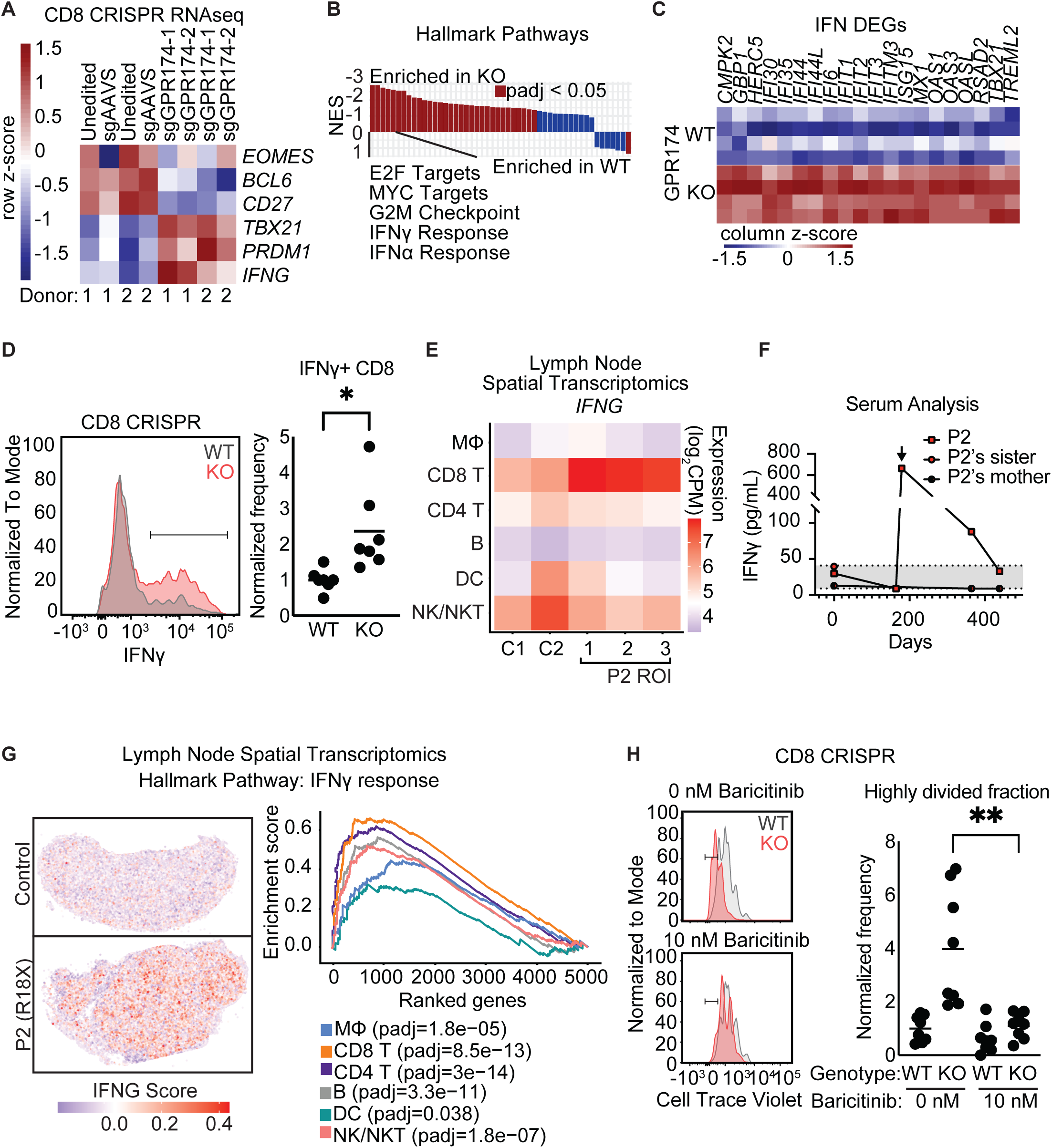
GPR174 deficiency and CD8 IFNγ signaling. A-C: RNA sequencing of *GPR174*-edited and control cells after 4 days of CD3/CD28 stimulation with 10 µM lysoPS. A. Heatmap of selected key CD8 genes. B. Hallmark Pathway analysis comparing GPR174 WT and KO cells. NES = normalized enrichment score. C. Heatmap of IFN inducible genes that are differentially expressed in WT versus KO. D: Flow cytometry evaluation of *GPR174*-edited and control cells after 4 days of CD3/CD28 stimulation with 10 µM lysoPS. Cells were restimulated with phorbol 12- myristate 13-acetate (PMA) and ionomycin and analyzed for IFNγ.χσβαρλινε E: *IFNG* transcript expression (log_2_ counts per million) in immune subsets across control and P2 lymph node ROIs from Xenium spatial transcriptomics. F: IFNγ cytokine profile of longitudinal plasma samples from P2 and related carriers. Arrow denotes steroid-treated flare. G: Left - Density map of the hallmark IFNγ Response pathway overlaid on control (C1) and P2 lymph nodes. Right - Enrichment across P2 immune subsets for the MSigDB Hallmark IFNγ Response gene set. Adjusted p-values are displayed. H: CRISPR edited CD8 T cells stimulated with CD3/CD28 beads in the presence of 10 µM lysoPS for 4 days +/- 10 nM baricitinib. Left shows example histogram plots of CTV dilution and right shows summary normalized frequency data for the highly divided fraction. Pooled from 4 independent experiments with 2 different donors, t-test with Welch’s correction, **p<0.01.

In *GPR174*-edited (KO) CD8 T cells, we observed a trend towards increased proliferation relative to control-edited or unedited (WT) CD8 T cells in response to CD3/CD28 stimulation (Fig 4A). Proliferation of WT but not *GPR174*-edited CD8 T cells was consistently reduced by lysoPS (Fig 4A). The trend towards increased proliferation in the edited group compared to the WT group without exogenous lysoPS likely represents variable levels of lysoPS in the culture at baseline. Analysis of patient-derived naïve CD8 T cells compared to healthy donor(s) revealed a similar resistance to lysoPS-mediated suppression of CD3/CD28-induced proliferation as observed for the CRISPR knock out cells (Fig 4B). These results demonstrate a prominent suppressive effect of lysoPS on human CD8 T cell proliferation that is lost in GPR174-deficient cells.

We next analyzed CD4 T cells. In contrast to mouse CD4 T cells and human CD8 T cells, both patient-derived and GPR174-edited healthy donor CD4 T cells proliferated similarly to control CD4 T cells and addition of lysoPS did not alter their proliferation (Fig 4C). Patient P2-derived CD4 T cells also proliferated comparably to healthy donors (Fig 4D). High levels of interleukin-2 (IL2) have been implicated in skewing CD8 T cells towards a terminally differentiated phenotype and suggested to promote T_EMRA_ differentiation (Geginat et al., 2003; Lee et al., 2021; Ross and Cantrell, 2018). CD4 T cells are a major source of IL2, and we and others have previously demonstrated that GPR174 attenuates mouse CD4 T cell IL2 production (Barnes and Cyster, 2018; Shinjo et al., 2017). We therefore evaluated the effect of GPR174 on human CD4 T cell IL2.

Despite the lack of proliferative difference, CD4 T cell IL2 production was attenuated by lysoPS in a GPR174-dependent manner (Fig 4E). These findings suggest that GPR174 loss independently alters CD4 and CD8 T cell function in a cell-autonomous manner.

The lineage-specific defects may cooperate to promote CD8 effector T cell overaccumulation and broader immune dysregulation.

To assess the effect of GPR174 loss on B cell proliferation, we measured B cell division in response to BCR stimulation in the presence and absence of lysoPS. Naïve B cells collected from healthy donors and P2 proliferated similarly in the presence or absence of exogenous lysoPS (Supp Fig 5B). There was also no GPR174-dependent effect of lysoPS on BCR-stimulated proliferation in B cells that had undergone CRISPR-editing (Supp Fig 5C). While these findings suggest a defect in B cell regulation may not be a major determinant in disease pathogenesis, they do not eliminate a GPR174 role in B cells as findings in mouse suggest B cell GPR174 function can be context-specific (Wolf et al., 2022).

### GPR174 deficiency promotes CD8 IFNγ production

Given the CD8 T_EMRA_ expansion in patients’ peripheral blood, CD8 expansion in P2 lymph node, and the hyperproliferative phenotype of GPR174-deficient CD8 cells, we hypothesized a central role for CD8 dysfunction in the disease state associated with GPR174 deficiency. To understand the cell-intrinsic molecular contributions of CD8 T cells to the disease phenotype, we profiled the transcriptomes of the *GPR174*-edited CD8 cells. Concordant with the observation of expanded CD8 T_EMRA_ in the patients, we found that these cells expressed transcripts consistent with a more proliferative and terminally differentiated effector phenotype. Analysis of important CD8 transcription factors and markers revealed a program associated with a more differentiated phenotype: more *TBX21* (encoding TBET) and *PRDM1* (BLIMP1) and less *BCL6* and *CD27* (Fig 5A, Supp Table 4). Consistent with the proliferative phenotype, gene signatures associated with active proliferation were highly enriched in the *GPR174*- edited cells (Fig 5B). In addition, supporting an effector phenotype, *IFNG* (interferon γ, IFNγ) expression was elevated and accompanied by a gene expression signature suggestive of autocrine signaling (Fig 5A-C). Increased IFNγ production was confirmed by flow cytometry (Fig 5D). Together, these findings suggest a cell-intrinsic effect of GPR174 loss on CD8 T cell proliferation and differentiation.

To gain molecular insights into GPR174-deficient cells within tissue, we performed pseudobulk transcriptional analyses on the lymph node spatial transcriptomics dataset. We found that among assessed P2 immune subsets, *IFNG* transcripts were most concentrated in CD8 T cells (Fig 5E). In addition, leveraging longitudinal P2 plasma samples, we found that peripheral blood from P2 showed elevated IFNγ during a disease flare different from the one at the time of biopsy, supporting a consistent role for IFNγ in disease flares across time (Fig 5F). P2’s sister and mother did not have elevations in IFNγ (Fig 5F). Importantly, when we compared differentially expressed genes (DEGs) between P2 immune subsets and control counterpart cells, we found that across all immune cell subsets, P2 cells were consistently and significantly enriched in an IFNγ response signature (Fig 5G). These findings indicate that IFNγ is abundant in and sensed by GPR174-deficient diseased tissues.

Since IFNγ signals via JAK1/2, we hypothesized that JAK inhibition might attenuate the hyperproliferation of GPR174-deficient CD8 T cells. Baricitinib is a dual inhibitor of JAK1 and JAK2, effective in the treatment of severe COVID-19 and a variety of autoinflammatory states (Marconi et al., 2021). We found that treatment of the *GPR174* CRISPR-edited CD8 T cells with baricitinib attenuated the hyperproliferative phenotype, suggesting a contribution of autocrine JAK/STAT signaling (Fig 5H).

### GPR174 deletion in mice expands virus-specific CD8 T cells in chronic infection

Since SPF GPR174-deficient mice are healthy and CD8 T_EMRA_ in humans are associated with cytomegalovirus (CMV) infection (Derhovanessian et al., 2011), we hypothesized that a chronic viral infection might reveal a role for murine GPR174 in host defense. Upon infection with lymphocytic choriomeningitis virus (LCMV) clone 13, GPR174 expression in the T cells of GPR174^+/TdTomato^ mice, as observed by the TdTomato knock-in reporter (Barnes et al., 2015), was down-regulated in the acute phase of infection (day 7) and restored in the chronic phase (day 21; Fig 6A). We therefore queried a regulatory role for GPR174 in the chronic phase of LCMV clone 13 infection. To test for CD8 T cell-intrinsic effects, we generated mixed chimeras with a 4:1 ratio of WT (CD45.1/2) to GPR174 WT or KO (CD45.2) bone marrow (Fig 6B). At 21 days post-infection, we found that the total CD8 ratio was similar between the WT/WT and WT/KO mixes, as was the ratio in the CD8+CD44+PD1+ population, which expresses GPR174 based on analysis of the TdTomato reporter (Fig 6C). However, KO cells were enriched in the virus-specific gp33+CD44+PD1+ CD8 T cell pool (Fig 6C).

**Figure 6:**
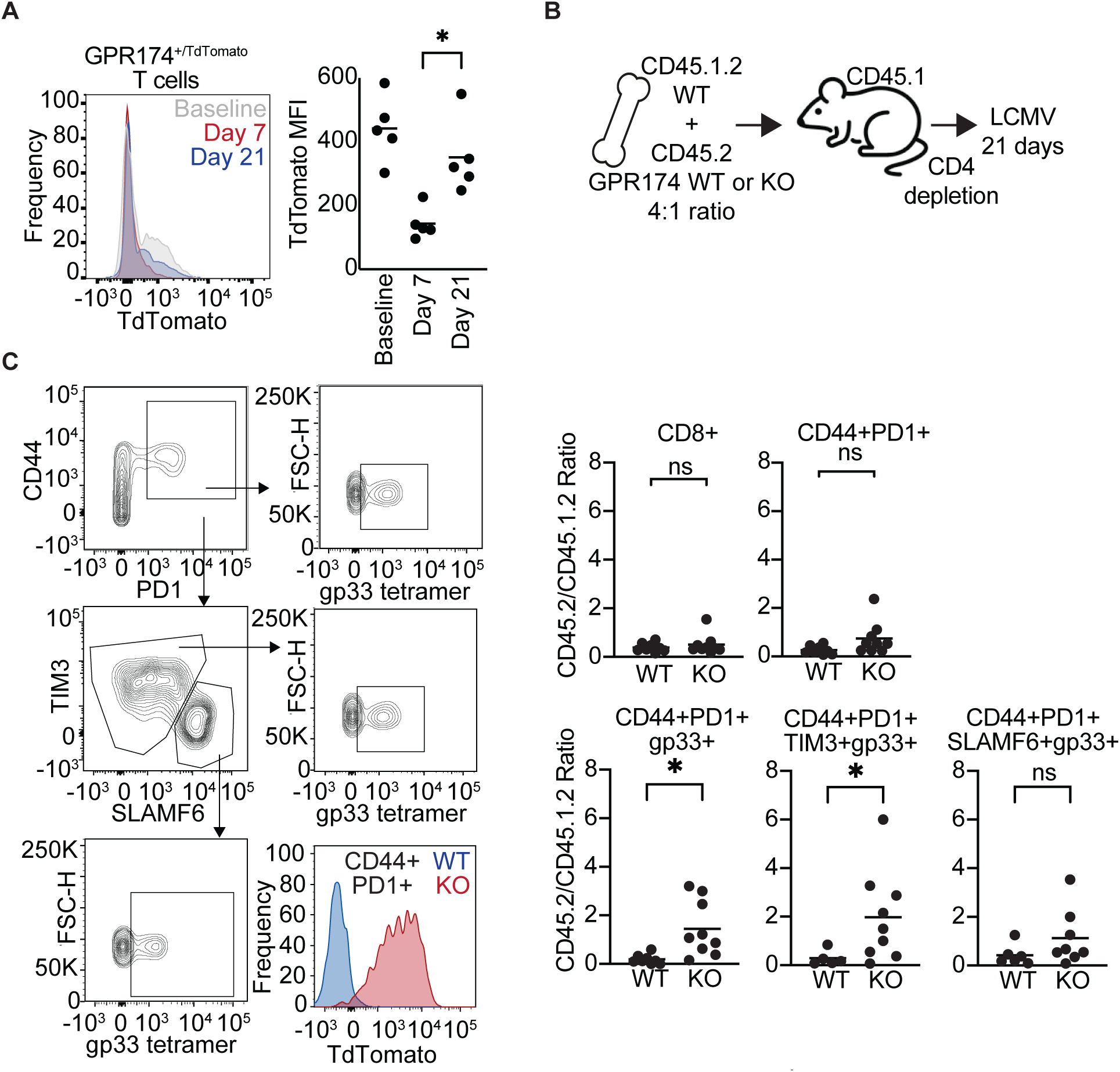
LCMV Clone 13 infection of GPR174-deficient mice. A: Flow cytometry analysis of spleen T cells from GPR174 TdTomato reporter heterozygous mice at day 7 and 21 after LCMV clone 13 infection. t-test with Welch’s correction, *p<0.05. B: Schematic showing CD45.1.2 WT and CD45.2 GPR174 WT or KO mixed bone marrow chimeras were generated at a 4:1 ratio, LCMV clone 13 infected, and analyzed at 21 days. C: Flow cytometry analysis of spleen CD8 T cells from day 21 LCMV clone 13 infected mice for the indicated markers. Graphs on right show ratio of test CD45.2 (WT or KO) cells over CD45.1.2 (WT) cells of indicated types. Each point is from an independent mouse, and data are pooled from 3 experiments. t-test with Welch’s correction, *p<0.05.

This difference was also present within the TIM3+ exhausted subset (Fig 6C). The SLAMF6+ T_pex_ pool trended in the same direction, but the increase did not reach statistical significance. These observations suggest a role of GPR174 in attenuating the antigen-specific CD8 T cell response to chronic viral infection.

## Discussion

The study of this cohort of individuals with GPR174 deficiency identifies a clinical phenotype characterized by lymphoid hyperplasia in the pattern of histiocytic necrotizing lymphadenitis and variable degrees of tissue autoimmunity, features shared with KFD. The disease-associated *GPR174* variants result in complete or partial loss of receptor expression, and complete or partial loss of function. In human T cells, GPR174 deficiency results in loss of lysoPS attenuation of IL2 production by CD4 T cells and of CD8 effector function in response to TCR stimulation. In chronic viral infection in mice, GPR174 loss favors the accumulation of virus-specific CD8 T cells. Together, these findings implicate GPR174 in attenuating antigen-specific T cell responses, the loss of which is associated with an increased risk of lymphadenopathy and autoimmunity (Fig 7).

**Figure 7:**
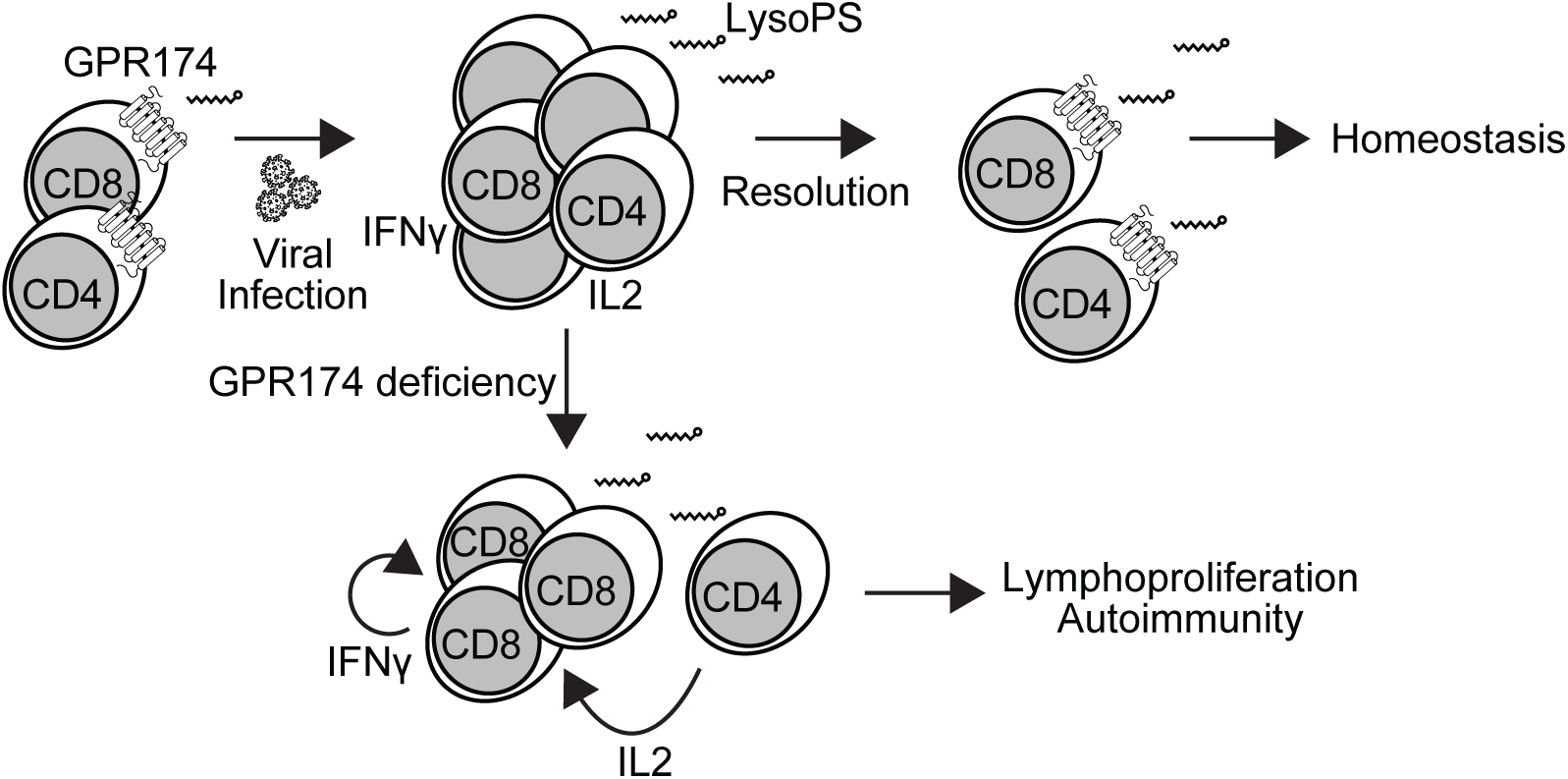
Summary figure. Model diagram showing naïve antigen specific T cells expressing GPR174 and being exposed to viral antigen. GPR174 is downregulated early (week 1) in T cell activation and then re-expressed at later stages. Activated CD4 cells provide IL2 that enhances CD8 T cell proliferation. GPR174 and lysoPS contribute to restraining CD4 IL2 production and CD8 expansion and IFNγ production, promoting immune response resolution. In the GPR174-deficient setting, this restraint on CD4 IL2, CD8 proliferation and IFNγ production is missing, predisposing to lymphoproliferation and autoimmunity.

Our data support a central role for CD8 T cells in the inflammatory response of our patients. These individuals have an expanded CD8 T_EMRA_ population *in vivo*, and we demonstrate that TCR-stimulated GPR174-deficient CD8 T cells hyperproliferate *in vitro*. CD8 T_EMRA_ populations expand in the presence of chronic or recurrent infections as well as in several autoimmune diseases, including autoimmune thrombocytopenia (Derhovanessian et al., 2011; Hamann et al., 1997; Malik et al., 2023; Xiong et al., 2023). While malignant clones were not detected in our patients, who were carefully evaluated for cancer, we do not rule out oligoclonal expansions in response to infection. The expanded CD8 population in GPR174-deficiency may contribute to a dysregulated response to infection (Hamann et al., 1997). This may parallel other IEIs triggered by infection, such as SAP deficiency (X-linked lymphoproliferative disease) (Purtilo et al., 1975). The role of a viral trigger is supported by the observation of severe disease in the index patient after COVID-19 infection, the virus-associated flares in some of the patients, and the over-expansion of virus-specific GPR174 KO cells in comparison to WT cells in the mouse LCMV clone 13 model. While there is not a defined mouse equivalent to human CD8 T_EMRA_ (Masopust et al., 2025), the CD44+PD1+TIM3+ CD8 population affected by GPR174 deficiency in the LCMV clone 13 analysis is a similarly highly differentiated state.

The finding that GPR174 attenuates CD8 effector responses is in alignment with previous studies of cAMP and G_s_-coupled GPCRs, such as prostaglandin receptors (EP2, EP4), catecholamine receptors (ADRB2), and adenosine receptors (A2AR), which suppress CD8 T cell responses (Chen et al., 2015; Globig et al., 2023; Stagg and Gutkind, 2025). Further investigation will be necessary to understand how chronic viral infections like CMV interact with GPR174 deficiency, as well as how the activities of GPR174 coordinate with other G_s_-coupled receptors to regulate CD8 expansion and effector function. Specificity in GPCR activity could be beneficial in therapeutic strategies to restrain or enhance the activity of T cell subsets in specialized contexts such as autoimmunity, infection, or cancer.

The full clinical phenotype of GPR174 deficiency is likely to be a combinatorial effect of receptor loss on multiple lymphocyte populations with a prominent role for T cells. While the heterozygous carriers related to P2 have an expanded CD8 T_EMRA_ population and experience recurrent febrile episodes, they lack the other manifestations such as lymphadenopathy and autoimmune cytopenias. The heterozygous carriers related to the other patients in the cohort have not developed clinically significant lymphadenopathy, autoimmunity, or infectious susceptibility. Other GPR174-regulated lymphocyte populations, such as CD4 T cells that produce IL2, may contribute to the phenotypic difference between the carriers and patients. Additional lymphocyte subsets may also play a role within tissues that are not easily sampled in the patient cohort. For example, in mice, the most prominent T_reg_ expansions associated with GPR174 loss are in the thymus and colon, which are not sites of routine sample collection in humans. Furthermore, the role of GPR174 in regulating B cell function *in vivo* remains elusive. There are some differences between our human and mouse studies - such as the observations that human CD8 T cells express more GPR174 than CD4 T cells, and that human CD4 T cells do not hyperproliferate in the absence of GPR174 – but further studies of mouse models will remain important for efforts to delineate the function of GPR174 in cell types and tissues not easily accessible in humans.

The efficacy of T cell targeted agents such as mycophenolate and sirolimus in alleviating inflammatory episodes in two of the patients further support a key role for T cells in the pathogenesis of the clinical disease. Recent investigations have demonstrated efficacy of JAK inhibition in APS-1, attenuating the role of T cells and IFNγ in that disease process (Oikonomou et al., 2024). The observation that the *in vitro* hyperproliferation of GPR174-deficient CD8 T cells is sensitive to JAK inhibition suggests a potential, targeted therapeutic strategy for individuals with GPR174 deficiency with the possibility to extend this approach to recurrent KFD.

In addition to the gene metrics suggesting that GPR174 does not undergo purifying selection, several observations indicate that GPR174-related lymphoproliferative disease likely displays incomplete penetrance and manifests in the setting of a relevant environmental stimulus and potentially additional genetic susceptibilities. While we do not have complete clinical data for the individuals in public databases with the R53X and R302W variants, the relatively high MAF of the R302W variant, together with a presumed absence of overt disease in most individuals, may indicate that additional genetic and environmental factors, such as viral infection, are required for disease to emerge. This may also explain the presence of other *GPR174* variants noted within public databases that may influence receptor function. In addition, there is heterogeneity in the clinical phenotype, and the degree of signaling defect does not correlate with the severity of the clinical disease. Variability in penetrance and expressivity is relatively common in inborn errors of immune dysregulation, including examples like CTLA4 haploinsufficiency (Schwab et al., 2018). Varying triggers may contribute to the specific sites of lymphoproliferation noted in different individuals, though it is also possible that generalized lymphadenopathy would have been detected in all patients with comprehensive imaging studies.

Morphologically, the lymph node biopsies available for review from this cohort resemble those of KFD. The pathogenesis of KFD is unclear. A viral trigger has been postulated but attempts to identify a unifying viral agent have yielded inconsistent results (Dominguez et al., 2003; Hollingsworth et al., 1994; Mahajan et al., 2023; Nelson et al., 2021). Within the lymph node, it has been proposed that type I interferon may promote cytotoxic CD8 T cell activity, resulting in tissue necrosis, as well as a T_H_1 response that involves the secretion of cytokines like IFNγ (Mahajan et al., 2023; Ohshima et al., 1998). In turn, IFNγ may contribute to the histiocyte/macrophage aggregates within the lymph node. The finding of an expanded CD8 T_EMRA_ population in the peripheral blood of GPR174-deficient individuals, a subset known for its cytotoxic activity, along with the expanded CD8 and macrophage populations in the patient lymph node, lends further support for an etiological connection between GPR174-deficiency and KFD. In a recent cohort of KFD patients, three clinical subtypes were described – one characterized by a higher incidence of fever, predominantly male, younger, and with a higher recurrence rate (Yu et al., 2025). Sequencing of this population may identify additional individuals with GPR174 deficiency or related genetic dispositions.

Identification of monogenic forms of KFD will facilitate further mechanistic studies of this disease entity, which may be due to a combination of genetic predisposition along with exposure to diverse potential viral triggers.

There is a growing collection of evidence linking viral infections and autoimmune diseases such as multiple sclerosis and systemic lupus erythematosus (Kim et al., 2026; Thomas et al., 2026; Wang et al., 2026; Younis et al., 2025). Inborn errors of immunity linking infectious susceptibility and autoimmunity such as GPR174 deficiency provide an important opportunity to investigate how these clinical conditions are connected.

Receptors like GPR174 may attenuate inflammation after an infectious insult, limiting tissue damage, cytokine production, and autoantigen exposure, for example. Future studies focused on the autoimmune propensity of GPR174 loss will be necessary to delineate the mechanisms. Such investigations into the regulation that differentiates protective from pathogenic lymphocyte responses has the potential to inform improved immunomodulatory therapies that permit protective immunity while suppressing damaging inflammation.

## Supplementary Figure Legends

Supplementary Figure 1: Further description of GPR174-deficient patients

A: Hematoxylin & eosin stain of a second cervical lymph node biopsy of P2. White arrowheads indicate lymphocytic infiltrate. Asterisks indicate B cell follicles. Scale bar = 100 µm.

B: CT scan of the chest of P6 demonstrating granulomatous-lymphocytic interstitial lung disease. Lung nodule biopsy revealed noncaseating granulomas.

Supplementary Figure 2: Schematic of reporter inserted via landing pad technology

A: GPR174 expression is inducible by doxycycline. An insulator prevents read-through to the GFP reporter. Puromycin (PuroR) and inducible caspase 9 (iCasp9) allow selection of cells with a recombination event inserting the GPR174 gene and CRE reporter.

Supplementary Figure 3: Further characterization of GPR174-deficient patient samples

A: Flow cytometry analysis of PBMCs collected from GPR174-deficient patients and carrier relatives compared to healthy controls. Each datapoint represents a unique individual. Group averages indicate median values. t-test with Welch’s correction, **p<0.01.

B: scRNAseq analysis of healthy female donor lymphocytes (Domínguez Conde et al., 2022; Ke et al., 2025; Xu et al., 2023) and female R18X carrier lymphocytes. Example genes selected based on SNP availability. Insufficient B and NK cells were captured in the carrier samples for analysis.

C: Analysis of allelic expression of *GPR174* in indicated cell types from single-cell datasets of carriers of the R18X variant (CA1, CA2) and a healthy donor (HD).

Supplementary Figure 4: Further characterization of GPR174-deficient patient lymph nodes

A: Cervical lymph node biopsies of P2 compared to a control reactive lymph node immunohistochemically stained to detect CD8 or Ki67. The samples were stained at different times; this analysis allows cell distribution but not staining intensity to be compared. Scale bar = 100 µm.

B: Hematoxylin & eosin staining of P2 biopsy #2 and two benign reactive follicular hyperplasia lymph node sections from pediatric male controls. Rectangles indicate approximate locations of regions of interest (ROIs) used for spatial transcriptomics analyses.

C: Annotated UMAP of cells contained within lymph node ROIs. Mϕ = macrophage. Mϕ/B and Mϕ/T indicate clusters where close approximation precludes resolution of macrophages from neighboring B and T cells. Due to overlapping transcriptional profiles, NK/NKT cells were retained as a single cluster.

D: Expression of key transcripts used to define cell types within ROIs for annotation.

Supplementary Figure 5: Characterization of GPR174-deficient B cells and TIDE analysis of CRISPR edits

A: TIDE analysis of B cell, CD4, and CD8 CRISPR edits.

B: Left – Experimental scheme: Naïve B cells collected from healthy donors (HDs) or a patient carrying the R18X variant (P2) were stimulated with αBCR, CD40L, IL-21, and IL-4 +/- 18uM lysoPS for 4 days. Center – Histogram plot shows example CTV dilution by stimulated healthy donor or P2 B cells. Right – Summary graph of lysoPS-mediated fold change in proliferation index from four HDs, including one same-day control as P2. C: Left – Experimental scheme: Naïve B cells were edited using sgRNAs targeting *GPR174* or *AAVS1*, then stimulated with αBCR, CD40L, IL-21, and IL-4 +/- 18uM lysoPS for 4 days. Center – Histogram plot shows example CTV dilution by stimulated healthy donor or P2 B cells. Right – Summary graph of lysoPS-mediated fold change in proliferation index from independent experiments with 3 donors.

Supplementary Table 1: Patient vignettes

Supplementary Table 2: R18X carrier scRNAseq supporting materials Cell assignments used for Supp Fig 3B-C CA1_Alleles, CA2_Alleles, HD_Alleles: X inactivation assignments Allelic_balance: Counts used to calculate Supp Figure 3C

Supplementary Table 3: Spatial transcriptomics supporting materials Cell_type_counts: Count data for Figure 3C IFNG_expression: Count data for Figure 5E IFN_GSEA: Enrichment scores corresponding to Figure 5G

Supplementary Table 4: CRISPR CD8 transcriptomics supporting materials Normalized_counts: Count data for Figure 5A LysoPS_WTvKO_DEGs: Differentially expressed genes used for Figure 5B/C GSEA_ranked_pathways: Full list of pathways in Figure 5B (left to right)

## Methods

### Regulatory compliance

Human samples were collected as part of routine clinical care or for research purposes with informed consent under the following protocols approved by the relevant ethics committees: Newcastle and North Tyneside Research Ethics Committee 1, REC ref 20/NE/0044. Children’s Hospital of Philadelphia, University of Pennsylvania Institutional Review Board, Protocol #21-018620 and 15-012265.

“Liguria CER” (Ethical Committee of Liguria Region) “Registry No.: 399/2021 - DB id 11649” (for Istituto G Gaslini in Genoa).

Animal experiments conformed to ethical principles and guidelines approved by the UCSF Institutional Animal Care and Use Committee.

### Genomic analysis

#### P1 Genome Sequencing

Genomics England Limited 100K Genome Project(The National Genomic Research Library, Genomics England (2024). https://doi.org/10.6084/m9.figshare.4530893) Immune Disorder Participant. Version 9 of Genomics England 100K Genome Project data was queried with a bespoke pipeline to identify potentially pathogenic variants that could be the cause of novel inborn errors of immunity (IEI). Rare, predicted damaging single nucleotide variants (SNVs) and short insertion/deletions (indels) from the GRCh38 aligned germline whole genome sequences of 940 immune disorder patients were compared with the germline variants from 15,238 cancer patients and separately the variants from 32,834 relatives of rare disease participants that did not have a recognised IEI. Variants with a minor allele frequency <0.001 that were predicted to cause high impact, loss of function, alterations (e.g. frameshift, stop-gain, canonical splice-site) in all protein coding genes (Ensembl release 94) that have not previously been shown to cause an IEI were extracted and counted. The per participant damaging variant counts were compared between immune patients and non-immune participant cohorts for each gene. The novel, hemizygous GPR174 p.Arg18Ter variant was the only identified rare, high impact hemizygous variant in GPR174 amongst the 940 immune patients and no such GPR174 variants were identified in the cancer germline or non-immune participant cohorts. Analysis pipeline scripts used to extract SNV/indel calls are as described previously(Shoemark et al., 2022) and available at https://github.com/Helgriff/Gel-rare-pipe.

#### P2 Genome Sequencing

Trio exome sequencing for patient P2 and his parents was performed via GeneDx. Briefly, after genomic DNA extraction, coding regions were captured using a proprietary system and paired-end sequenced on an Illumina platform. Reads were aligned to GRCh37/UCSC hg19, achieving a mean coverage depth of 204X. Variants as well as deletions and duplications with >3 exons were identified using XomeAnalyzer with a quality threshold of 98.9 (Retterer et al., 2015).

#### P3 Genome Sequencing

DNA was extracted from blood samples collected in ethylene diamine tetraacetic acid-tubes. WGS library preparation, sequencing, and bioinformatics analysis were performed on DNA using standard protocols. Reads were aligned to the human reference genome GRCh37 and variants were called and annotated using center-specific bioinformatics pipelines. Variant assessment incorporated population frequency data and computational prediction tools, including phyloP, SIFT, PolyPhen-2, FATHMM, CADD, and REVEL. The identified variant was queried in LOVD (https://databases.lovd.nl/shared/), ClinVar (https://www.ncbi.nlm.nih.gov/clinvar), and gnomAD (https://gnomad.broadinstitute.org/). The interpretation of sequence variants followed the American College of Medical Genetics and Genomics guidelines (Richards et al., 2015).

#### P4/5 Genome Sequencing

DNA was extracted from peripheral blood samples of the patients and healthy mother using QIAamp DNA Blood Midi kit (Qiagen, Germantown, MD, USA). The quality and quantity of DNA thus obtained were determined by Nanodrop. Whole-exome sequencing (WES) was performed on an Illumina NovaSeq 6000 platform, after exome capture using the Nextera Rapid Capture Expanded Exome Kit (Illumina Inc., San Diego, CA, USA). An average sequencing depth of 200× was achieved across all samples, with more than 99% of the coding regions covered at a minimum depth of 20×. Raw sequencing data (FASTQ files) underwent quality control using FastQC. Variant calling was carried out using an in-house pipeline (https://github.com/igg-bioinfo/diva.wes), with the GRCh38 (hg38) human genome assembly as the reference. Variants were annotated using Ensembl Variant Effect Predictor (VEP) v107, incorporating multiple annotations including allele frequencies from gnomAD v3.0, CADD pathogenicity scores, and clinical significance from the ClinVar database. For the identification of candidate germline variants, we retained variants that were either absent from gnomAD or had a minor allele frequency (gnomADg_AF) < 0.01. Segregation analysis was performed within the family in accordance with the observed phenotype.

#### P6 Genome Sequencing

Exome sequencing was performed using standard exome capture and sequencing protocols. Raw data were analyzed using Varfeed and variants were annotated and prioritized with Varvis, focusing on protein-altering variants and their clinical relevance across all inheritance models. Only variants in OMIM disease-associated genes are reported, with rare exceptions supported by strong external evidence; variants without clear clinical relevance were not reported.

#### Public database analysis

Publicly available variant details for the GPR174 gene were downloaded from GnomAD v4.1 (https://gnomad.broadinstitute.org/gene/ENSG00000147138?dataset=gnomad_r4) & Topmed freeze10 (https://bravo.sph.umich.edu/gene.html?id=GPR174) databases.

Variants were filtered to those that had been seen in hemizygous form in at least one male individual and that had a nonsynonymous effect prediction. The CADD score for each hemizygous variant was plotted against its reported minor allele frequency (MAF). The four hemizygous variants identified in the patient cohort are labeled in red.

### Expression vectors

Human *GPR174* was PCR amplified from tonsil cDNA and an N-terminal prolactin signal peptide and OX56 epitope tag was added (Ishii et al., 1993; Lu et al., 2019). The PCR insert was introduced to the pQEF-Ceru-T2A vector (Yang and Allen, 2018) by Gibson assembly (NEB, E2611S). Site-direct mutagenesis using the primers in the table below and Phusion high fidelity DNA polymerase (NEB, M0530L) was performed to generate the patient-derived GPR174 variants.

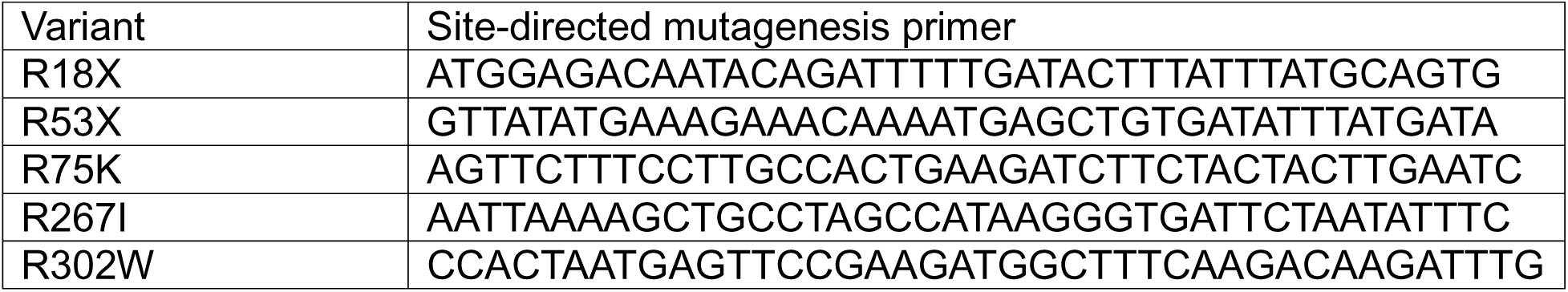

### cAMP reporter

Reporter cells were generated as previously described (Howard et al., 2025). Briefly, GPR174 variants were PCR amplified from the pQEF vector and cloned by Gibson assembly into pLP_mod_mNeonGreen11_P2A_puroR_CRE-GFP (gift from Willow Coyote-Maestas, Addgene # 229227), which carries a destabilized GFP reporter under a cAMP responsive element promoter. This vector was transfected with Lipofectamine 3000 (Thermo Fisher Scientific) along with BxB1 recombinase (pCAG-NLS-BxB1, gift from Pawel Pelczar, Addgene #51271) into HEK293T-LLP-iCasp9-Blast cells, which have a lentiviral landing pad to facilitate single copy, single locus integration (Matreyek et al., 2020). Gene expression from a tetracycline-responsive promoter was induced with 2 µg/mL doxycycline (Millipore Sigma, D9891). This expresses a split rapamycin analog-dimerizable caspase 9 in cells without a recombination event and a puromycin resistance gene from cells with a recombination event. Reporter cells were selected for two days in 10 nM AP1903 (MedChemExpress, HY-16046), which causes dimerization of caspase 9 and cell death, followed by two days in 2 µg/mL puromycin (InVivoGen, ant-pr-1). Cells were maintained in tetracycline-free D10 media composed of DMEM (Corning, MT10013CV), 10% Fetal Bovine Serum (FBS, Omega Scientific, FB-02), 1% penicillin-streptomycin (UCSF Media Core, CCFGK004), 1% sodium pyruvate (Gibco, 11360070).

For receptor expression and reporter assays, the reporter cells were plated at 20,000 cells per well in 96 well plates overnight in D10 media. Receptor expression was induced with 100 ng/mL doxycycline for 24 hours prior to assay collection. This concentration was selected based on titration to reliably induce but not saturate receptor expression. For measurement of the transcriptional reporter, cells were treated with 10 µM trimethoprim (MedChemExpress, HY-B0510) for 4 hours to stabilize the GFP prior to assay collection.

### Western blot

Receptor expression was induced with 100 ng/mL doxycycline for 24 hours prior to sample collection. 5e6 cells were lysed for 1 hour on ice in GPCR lysis buffer: 0.5% Brij 35 (Santa Cruz sc-280628), 0.5% NP40 (Thermo Fisher, 85124), 150 mM NaCl (Fisher, S671-10), 10 mM Tris-HCl pH 7.4 (Fisher, BP152-5 & A144-212), protease inhibitor (Sigma, 11836170001). To enrich GPR174, lysates were incubated with 10 µg anti- OX56 antibody (BioXCell, custom) overnight and then with 30 µL magnetic Protein G beads (NEB, S1430S) for 1 hour. Protein denaturation was performed with lithium dodecyl sulfate sample buffer (NuPAGE, NP0007) supplemented with β-mercaptoethanol (Biorad, 1610710) at room temperature for 15 minutes. Proteins were separated on 4-12% Bis-Tris SDS-PAGE gels (Invitrogen, NP0323) and transferred to a methanol-activated PVDF membrane. Western blot protein detection was performed using anti-OX56-biotin (1:1000), anti-tubulin (CST, 2128T, 1:1000) primary antibodies, IRDye 800CW goat anti-rabbit IgG (LICOR, 926-68079), and IRDye 680RD streptavidin (LICOR, 926-68079) antibodies. Images were acquired with the LICOR Odyssey CLx system and quantified with ImageStudio.

### Flow cytometry

Cells were stained in 96 well round-bottom plates on ice in PBS with 2% newborn calf serum (Fisher, 16010142) and 1 mM EDTA (Fisher, BP2482-500) for 20 minutes. Antibodies are listed below. LCMV-specific cells were identified by tetramer staining with H-2D(b) LCMV gp 33-41 KAVYNFATC AlexaFluor-647 (NIH Tetramer Core Facility) for 30 min at room temperature prior to adding additional antibodies. For cytokine stains, cells were re-stimulated with 50 ng/mL phorbol 12-myristate 13-acetate (Sigma, P1585) and 750 ng/mL ionomycin (Sigma, 10634) in the presence of Golgistop protein transport inhibitor (Fisher, 554724) for 2 hours prior to staining. Viability was determined using Fixable Viability Dye eFluor780. Fixation and permeabilization for intracellular stains were accomplished with BD Cytofix/Cytoperm (Fisher, 554722) and BD Perm/Wash (Fisher, 554723) according to manufacturer’s instructions. Data were collected on a FACSymphony A1 cytometer. Post-acquisition analysis was performed with FlowJo (v10-11).

| Antigen, Clone | Conjugate | Source | Dilution |
| --- | --- | --- | --- |
| Human CD8a, RPA-T8 | BV785 | Biolegend | 1:100 |
| Human CD4, OKT4 | APC | Biolegend | 1:200 |
| Human CD3, OKT3 | BV605 | Biolegend | 1:200 |
| Human CD3, UCHT1 | FITC | Biolegend | 1:200 |
| Human CD19, HIB19 | PE-Cy7 | Biolegend | 1:200 |
| Human CD14, HCD14 | PE | Biolegend | 1:100 |
| Human IFNγ, B27 | PE-Cy7 | Biolegend | 1:200 |
| Mouse LY108, 330-AJ | Pacific Blue | Biolegend | 1:100 |
| Mouse CD45.1, A20 | BV605 | Biolegend | 1:200 |
| Mouse TIM3, RMT3-23 | BV711 | Biolegend | 1:100 |
| Mouse CD44, IM7 | BV785 | Biolegend | 1:200 |
| Mouse CD45.2, 104 | FITC | Biolegend | 1:200 |
| Mouse CD8a, 53-6.7 | PerCP-Cy5.5 | Tonbo | 1:100 |
| Mouse PD1, RMP1-30 | PE-Cy7 | Biolegend | 1:100 |
| Anti-OX56 (Rat CD43) | Biotin | BioXcell, in-house biotinylation | 1:100 |

### Patient Immunophenotyping

Patient and age/sex-matched healthy donor peripheral blood mononuclear cells (PBMCs) samples were isolated using Ficoll-Paque PLUS density gradient centrifugation (Cytiva). Cells were stained in 1.5ml microcentrifuge tubes at 4°C in PBS with 0.5% BSA (Cell Signaling, 9998S) for 30 minutes. Antibodies are listed below.

Fixation and permeabilization for intracellular stains were done with Foxp3/ Transcription Factor Staining Buffer Set (eBioscience, 00-5523-00) according to manufacturer’s instructions. Immunophenotyping data were captured by an Aurora Spectral Analyzer (CyTek Biosciences) and analyzed with FlowJo software (TreeStar).

| Antigen | Clone | Conjugate | Source | Dilution |
| --- | --- | --- | --- | --- |
| Fixable Viability Dye | — | eFluor 780 | eBioscience | 1:800 |
| Human CD1c | L161 | Super Bright 436 | eBioscience | 1:25 |
| Human CD3 | UCHT1 | Pac Orange | Thermofisher | 1:25 |
| Human CD3 | UCHT1 | AF700 | Biolegend | 1:100 |
| Human CD4 | OKT4 | APC/Cy7 | Biolegend | 1:100 |
| Human CD4 | OKT4 | AF700 | Biolegend | 1:50 |
| Human CD4 | OKT4 | FITC | Biolegend | 1:100 |
| Human CD8 | RPA-T8 | BUV395 | BD OptiBuild | 1:50-100 |
| Human CD10 | HI10a | PE-Cy7 | Biolegend | 1.5:50 |
| Human CD11c | 3.9 | BV650 | Biolegend | 3:50 |
| Human CD14 | HCD14 | Pacific Blue | Biolegend | 1:50 |
| Human CD14 | M5E2 | BUV496 | BD OptiBuild | 1:50 |
| Human CD15s | CSLEX1 | BV711 | BD Bioscience | 1.5:50 |
| Human CD16 | B73.1 | FITC | Biolegend | 1:50 |
| Human CD16 | 3G8 | BUV496 | BD OptiBuild | 1:50 |
| Human CD19 | HIB19 | BUV496 | BD OptiBuild | 1:50 |
| Human CD20 | 2H7 | AF700 | Biolegend | 3:50 |
| Human CD21 | B-ly4 | BUV563 | BD OptiBuild | 1:50 |
| Human CD21 | B-ly4 | PE | BD Bioscience | 1:50 |
| Human CD24 | MI5 | BV785 | Biolegend | 2:25 |
| Human CD25 | 2A3 | BUV737 | BD OptiBuild | 1:50 |
| Human CD25 | M-A251 | PeDazzle 594 | Biolegend | 1:50 |
| Human CD25 | 3A3 | BUV737 | BD OptiBuild | 1:100 |
| Human CD25 | BC96 | PE | Biolegend | 1:100 |
| Human CD27 | O323 | BV510 | Biolegend | 1:12.5-50 |
| Human CD27 | O323 | BV570 | Biolegend | 1:50 |
| Human CD28 | CD28.2 | Super Bright 436 | eBioscience | 1:20 |
| Human CD38 | HIT2 | APC | Biolegend | 1.5:50 |
| Human CD38 | HIT2 | AF488 | Biolegend | 1.5:50 |
| Human CD39 | A1 | BV510 | Biolegend | 1:50 |
| Human CD45RA | HI100 | BUV805 | Biolegend | 1:100 |
| Human CD45RO | UCHL1 | AF700 | Biolegend | 1:20 |
| Human CD45RO | UCHL1 | BUV737 | BD OptiBuild | 1:50 |
| Human CD56 | HCD56 | BV510 | Biolegend | 1.5:50 |
| Human CD57 | HNK-1 | PeDazzle 594 | Biolegend | 1:200 |
| Human CD62L | DREG-56 | BUV661 | BD OptiBuild | 1:50 |
| Human CD69 | FN50 | PE | Biolegend | 1:50 |
| Human CD69 | FN50 | Pe/Cy7 | Biolegend | 1.5:100 |
| Human CD86 | FUN-1 | BV750 | BD OptiBuild | 1:100 |
| Human CD95 (Fas) | DX2 | BV605 | Biolegend | 1:20 |
| Human CD95 (FASR) | DX2 | APC | Biolegend | 1.5:100 |
| Human CD117 | 104D2 | APC/Cy7 | Biolegend | 1.5:50 |
| Human CD127 | A019D5 | PerCP/Cy5.5 | Biolegend | 1:25 |
| Human CD127 | A019D5 | BV650 | Biolegend | 1:25 |
| Human CD134 (OX40) | ACT35 | BV711 | BD Bioscience | 1:50 |
| Human CD141 | M80 | APC | Biolegend | 1.5:50 |
| Human CD152 (CTLA-4) | BNI3 | BV786 | BD Bioscience | 1.5:50 |
| Human CD183 (CXCR3) | G025H7 | FITC | Biolegend | 1:50 |
| Human CD185 (CXCR5) | J252D4 | Pac Blue | Biolegend | 1:50 |
| Human CD196 (CCR6) | 11A9 | PE | BD Bioscience | 1:50 |
| Human CD197 (CCR7) | 3D12 | BV786 | BD Bioscience | 1:50 |
| Human CD268 (BAFF-R) | 11C1 | PE | Biolegend | 1:50 |
| Human CD272 (BTLA) | J168-540 | BV650 | BD OptiBuild | 1.5:50 |
| Human CD278 (ICOS) | ISA-3 | Super Bright 436 | eBioscience | 1:25 |
| Human CD279 (PD-1) | EH12.2H7 | PE/Cy7 | Biolegend | 1:50 |
| Human CD294 (CRTH2) | BM16 | eFluor660 | eBioscience | 3:50 |
| Human CD303 | 201A | PeCy7 | Biolegend | 1:50 |
| Human HLA-DR | G46-6 | BV750 | BD Bioscience | 1:50 |
| Human TCRa/b | IP26 | APC | Biolegend | 1:25 |
| Human TCRg/d | B1 | PeDazzle 594 | Biolegend | 1:50 |
| Human TCRg/d | B1 | PerCP/Cy5.5 | Biolegend | 1:25 |
| Human IgD | IA6-2 | AF700 | BD Bioscience | 2:25 |
| Human IgM | G20-127 | PerCP-Cy5.5 | BD Bioscience | 1:50 |
| Human Foxp3 | 150D | AF488 | Biolegend | 3:50 |
| Human Helios | 22F6 | PerCpCy5.5 | Biolegend | 3:50 |
| Human Ki67 | Ki67 | PE | Biolegend | 1.5:50 |
| Human Pu.1 | 7C6B05 | AF647 | Biolegend | 1:20 |
| Human T-bet | 4B10 | Pe/Dazzle 594 | Biolegend | 1:100 |
| Human TIGIT | A15153G | BV605 | Biolegend | 1:25 |

#### Cell type definitions

Central memory CD4: CD3+ TCRab+ CD4+ CD45RA- CCR7+

Effector memory CD4: CD3+ TCRab+ CD4+ CD45RA- CCR7-

Naïve CD4: CD3+ TCRab+ CD4+ CD45RA+ CCR7+

T_reg_s: CD3+ TCRab+ CD4+ FoxP3+ CD25+

Central memory CD8: CD3+ TCRab+ CD8+ CD45RA- CCR7+

Effector memory CD8: CD3+ TCRab+ CD8+ CD45RA- CCR7-

Naïve CD8: CD3+ TCRab+ CD8+ CD45RA+ CCR7+

T_EMRA_: CD3+ TCRab+ CD8+ CD45RA+ CCR7- CD27-

Naïve B cells: CD19+ CD27- IgD+

Memory B cells: CD19+ CD27+

Class-switched memory B cells: CD19+ CD27+

IgD- Plasmablasts: CD19+ CD27+

IgD- CD38hi DN2 B cells: CD19+ CD27- IgD- CD21- CD19hi

Dendritic cells: CD3- CD14- CD20- HLA-DR+

Plasmacytoid DCs: CD3- CD14- CD20- HLA-DR+ CD303+ CD11c-

Conventional DCs: CD3- CD14- CD20- HLA-DR+ CD303- CD11c+

NK cells: CD3- CD4- CD16+ CD56+

Classical monocytes: monocyte FSC/SSC gate, CD16- CD14+

Intermediate monocytes: monocyte FSC/SSC gate, CD16int CD14int

Non-classical monocytes: monocyte FSC/SSC gate, CD16+ CD14-

### Single-cell RNA sequencing

Cryopreserved PBMCs were thawed and sorted using a BD FACSAria II cytometer to enrich for live lymphocytes (viability dye+ cells and CD3-CD19-CD14+ cells were removed). Single-cell RNA sequencing was performed with the 10X 3’ GEM-X v4 kit. 10,933 – 18,889 cells were collected per sample and sequenced on Illumina NovaSeq X Plus. 9.2-11.6*10^8^ reads were collected per sample, yielding 48,656 – 88,012 mean reads per cell, and detecting 2526 - 2700 median genes per cell. Data were mapped to the GRCh38-2024-A genome using CellRanger 9.0.1.

Bam files generated from CellRanger were further processed as previously described for analysis with scLinaX (Tomofuji et al., 2024a). Briefly, X chromosome reads were extracted and processed using SAMtools v1.21 (Danecek et al.). SNPs were genotyped with Cellsnp-lite v1.2.3 (Huang and Huang, 2021) and annotated with ANNOVAR (2022) (Wang et al., 2010). Cell types were assigned based on a PBMC reference using Seurat v5.2.1 and Azimuth v0.5.0 (Hao et al., 2021). Cell quality was filtered based on mitochondrial content, transcript reads, and doublet detection using DoubletFinder (McGinnis et al., 2019). The active and inactive X chromosome were annotated using scLinaX v0.1.0 (Tomofuji et al., 2024a).

Cell type annotation was performed as follows using Seurat: T cells were identified by *CD3E* and *TRAC* expression. B, γδT, NK, and monocytes were excluded based on *CD79A*, *TRDC*, *NCR1*, and *LYZ* expression. T cells were clustered based on gene expression, and conventional CD8 cells were identified based on clusters that expressed both *CD8A* and *CD8B*. CD4 clusters were also identified for comparison. The CD8 clusters were then sub-clustered, and clusters were analyzed for markers of CD8 differentiation. Cluster 5 demonstrated the lowest expression of naïve-associated markers including *TCF7, CCR7, CD27, IL7R* with the highest expression of differentiation-associated genes *TBX21, PRDM1, GZMB,* and *PRF1*, consistent with the profile of CD8 T_EMRA_. Clusters 0-2 were designated CD8 Naïve/TCM based on high *CCR7* expression.

### Human T cell culture

Naïve CD4 and CD8 T cells were enriched from leukapheresed PBMCs (StemCell Technologies) using antibody-based magnetic bead negative enrichment (StemCell Technologies, 17555 & 17968) and maintained in X-VIVO 15 media (Lonza, 02-060Q) supplemented with 5% FBS, 50 mM β-mercaptoethanol (Gibco, 21985023), 10 mM N-Acetyl L-Cysteine (Sigma, A9165) as previously described (Shifrut et al., 2018). 5 ng/mL human IL7 (PeproTech, 200-07) was added for all naïve cultures. Cell culture maintenance and assays were performed in 96 well, flat-bottom suspension culture plates.

CRISPR editing was performed by activating T cells with CD3/CD28 beads at a ratio of 1 bead:10 cells (Thermo Fisher, 11132D), 300 U/mL human IL2 (PeproTech, 200-02), 5 ng/mL human IL7, and 5 ng/mL human IL15 (PeproTech, 210-15) for two days followed by nucleofection (Lonza, V4SP-3096) with ribonucleoprotein complexes of crRNA (IDT, Alt-R), tracrRNA (IDT, Alt-R), and Cas9 protein (University of California-Berkeley MacroLab). Cells were cultured in supplemented X-VIVO media with 300U/mL human IL2 for two days prior to further assessment.

For proliferation assays, T cells were rested overnight at 250,000 cells per well in supplemented XVIVO media without IL2. Cells were stained with CellTrace Violet (Invitrogen, C34557) for 10 minutes at room temperature for dye dilution assays, then plated at 20,000 cells per well and stimulated with CD3/CD28 beads with the indicated amount of 18:1 lysoPS (Avanti Polar Lipids, 858143P) in RPMI supplemented with 10% FBS, 1 mM HEPES (Fisher, MT25060), L-alanyl-L-glutamine dipeptide (Fisher, 35050061), 1% penicillin-streptomycin, and 55 nM β-mercaptoethanol. 10 nM baricitinib (MedChemExpress, HY15315) was added as indicated. Cells were collected at four days post-stimulation for analysis. In the graphs, the WT group includes unedited and safe-harbor targeted sgRNA samples, and the KO group includes two independent GPR174-targeted sgRNAs. To facilitate pooling from different experiments, values are normalized to the average of the unedited group treated with lysoPS in each experiment.

To assess editing efficiency, edited cells maintained in IL2 were collected at the time of post-stimulation analysis. Genomic DNA was collected by lysis in QuickExtract DNA Extraction Solution (Lucigen, VWR 76081-766) followed by PCR with flanking primers and analysis using TIDE (Brinkman et al., 2014). Editing efficiency is shown in Supp Fig 5A.

### B cell-editing proliferation assays

Naïve B cells were enriched from leukapheresed PBMCs using antibody-based magnetic bead negative enrichment (Biolegend, 480068). CRISPR editing was performed by nucleofection (Lonza, V4XP-3024) with ribonucleoprotein complexes of crRNA (IDT, Alt-R), tracrRNA (IDT, Alt-R) and Cas9 protein (IDT, Alt-R). Cells were cultured in ImmunoCultTM -XF B cell Base Medium (StemCell Technologies, 100-0646) supplemented with ImmunoCultTM -ACF Human B cell Cell Expansion Supplement (StemCell Technologies, 10974) for 24 hours prior to proliferation assays. B cells were stained with Tag-it Violet™ Proliferation and Cell Tracking Dye (Biolegend, 76806) for 8 minutes at 37C, then plated in a 96-well U bottom plate at 50,000 cells per well and stimulated with 2.5ug/ml AffiniPure® F(ab’)_2_ Fragment Goat Anti-Human IgA + IgG + IgM (H+L) (Jackson ImmunoResearch, 109-006-064), 25ng/ml IL-21 (R&D, 8879-IL- 010), 10ug/ml IL-4 (Peprotech, 200-04-5UG), and 5ug/ml CD40L (Biolegend, 591706), with the indicated amount of 18:1 lysoPS (Avanti Polar Lipids, 858143P) in RPMI supplemented with 10% FBS, 2mM L-Glutamine (Corning, 25-005-CI), 1% penicillin-streptomycin (Gibco, 15140-122), 1mM Sodium Pyruvate (Gibco, 11360-070), 10mM MEM NEAA (Gibco, 11140-050), 10mM HEPES (Gibco, 15630-080) and 1ug/ml rBaff (Biolegend, 559608). Cells were collected at four days post-stimulation for analysis. To assess editing efficiency, genomic DNA was extracted 24 hours post-nucleofection followed by PCR with flanking primers and analysis using TIDE.

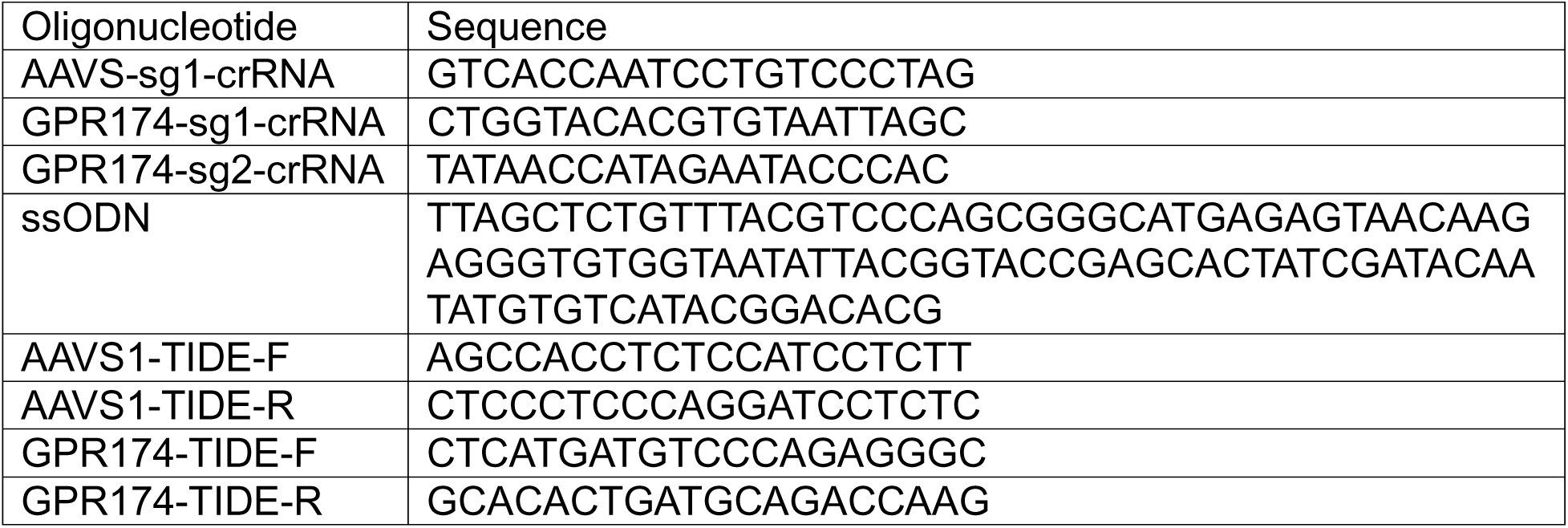

### Patient T cell proliferation assays

Naïve CD4 and CD8 T cells were sorted (CD4+ or CD8+, CD25-, CD45RA+, CD62L+) on an Aurora CS (CyTek Biosciences) following PBMC isolation. Cells were stained with Tag-it Violet™ Proliferation and Cell Tracking Dye (Biolegend, 76806) for 8 minutes at 37C, then plated in a 96-well U bottom plate at 10,000 cells per well and stimulated 1:1 with Dynabeads™ Human T-Activator CD3/CD28 (Gibco, 11131D) with the indicated amount of 18:1 lysoPS (Avanti Polar Lipids, 858143P) in RPMI supplemented with 10% FBS, 2mM L-Glutamine (Corning, 25-005-CI), 1% penicillin-streptomycin, 1mM Sodium Pyruvate (Gibco, 11360-070). Cells were collected at four days post-stimulation for analysis.

### Patient B cell proliferation assays

Naïve B cells were sorted (CD19+, CD21+, CD27-) on an Aurora CS (CyTek Biosciences) following PBMC isolation. Cells were stained with Tag-it Violet™ Proliferation and Cell Tracking Dye (Biolegend, 76806) for 8 minutes at 37C, then plated in a 96-well U bottom plate at 50,000 cells per well and stimulated with 2.5ug/ml AffiniPure® F(ab’)_2_ Fragment Goat Anti-Human IgA + IgG + IgM (H+L) (Jackson ImmunoResearch, 109-006-064), 25ng/ml IL-21 (R&D, 8879-IL-010), 10ug/ml IL-4 (Peprotech, 200-04-5UG), and 5ug/ml CD40L (Biolegend, 591706), with the indicated amount of 18:1 lysoPS (Avanti Polar Lipids, 858143P) in RPMI supplemented with 10% FBS, 2mM L-Glutamine (Corning, 25-005-CI), 1% penicillin-streptomycin (Gibco, 15140- 122), 1mM Sodium Pyruvate (Gibco, 11360-070), 10mM MEM NEAA (Gibco, 11140- 050), 10mM HEPES (Gibco, 15630-080) and 1ug/ml rBaff (Biolegend, 559608). Cells were collected at four days post-stimulation for analysis.

### ELISA

Supernatants were collected from proliferation cultures after four days of stimulation.

IL2 ELISA was performed using a human IL2 uncoated ELISA kit per manufacturer’s instructions (Fisher, 88-7025-22).

### RNA analysis

RNA was collected using a column-based kit (Qiagen, 74004) per manufacturer’s instructions. cDNA was generated with M-MLV reverse transcriptase (Invitrogen, 28025013). Quantitative polymerase chain reaction was performed with Power SYBR Green PCR Master Mix (Life Tech, 4367660). Library construction for RNA sequencing was performed with Ovation RNA-seq System V2 (NuGEN, 7102-A01) and Kapa hyper prep kit (Roche, 07962347001) and sequencing was performed with Illumina NovaSeq X Plus.

Read quality was checked using FastQC v0.12.1 (https://www.bioinformatics.babraham.ac.uk/projects/fastqc/) and aligned to the hg38 genome using STAR aligner v2.7.11b (Dobin et al., 2013), using standard settings for paired reads. Uniquely mapped reads were assigned to annotated genes with HTSeq v2.0.5 (Putri et al., 2022) with standard settings. Read counts were normalized by library size, and differential gene expression analysis based on a negative binomial distribution was performed using DESeq2 v1.44.0.(Love et al., 2014) Gene set enrichment analysis was performed with fgsea v1.30.0 (Korotkevich et al., 2021).

### Spatial transcriptomic analysis

Five regions of interest (ROIs) were identified in 5-μm FFPE lymph node sections obtained from two control subjects (C1, 6-10 year old male, cervical non-malignant reactive lymphadenopathy, one ROI; C2, 11-15 year old male, cervical non-malignant reactive lymphadenopathy, one ROI) and one GPR174-deficient patient (P2, cervical proliferative-phase Kikuchi-Fujimoto disease, three ROIs). ROIs were selected to capture non-contiguous regions containing multiple germinal centers while avoiding fibrotic areas. Tissue sections were maintained in a desiccator until use. Duplicate tissue sections representing each ROI were distributed across two Xenium slides and processed in parallel as technical replicates. ROIs were profiled using the 10x Genomics Xenium In Situ platform with the Xenium Human 5K panel according to the manufacturer’s protocols (CG000580 Rev E; CG000760).

Following image acquisition and transcript decoding using the Xenium onboard analysis pipeline, downstream analyses were performed in R (v4.4.0) using the Seurat package (v5.5.0) (Hao et al., 2024). Transcript detections were filtered to retain molecules with Xenium quality values (QV) ≥ 20. Per-cell quality control metrics, including total transcript counts and detected genes, were calculated, and cells with fewer than 50 transcripts or fewer than 45 detected genes were excluded from downstream analyses. Following filtering, approximately 1.5 million, 3.1 million, and 2.7 million cells were retained from samples C1, C2, and P2, respectively. Median transcript counts per cell ranged from 113 to 383 depending on the ROI.

To enable scalable analysis of the large spatial transcriptomic dataset, cells were subsampled using the Seurat SketchData workflow, which preserves biological complexity through proportional representation of both common and rare cell populations. Prior to sketching, RNA counts were normalized using Seurat’s LogNormalize method, in which gene expression values for each cell are normalized by total expression, multiplied by a scale factor of 10,000, and log transformed. Variable features were identified, and a sketch consisting of 20,000 representative cells was generated using leverage score sampling. The sketch assay was subsequently normalized, highly variable features were identified, and expression values were centered and variance-scaled using Seurat’s ScaleData function without regression of covariates. Principal component analysis (PCA) was performed on the scaled sketch data, and donor-specific effects were corrected using Harmony integration. Based on inspection of the PCA structure and elbow plot, the first 10 principal components were used for neighborhood graph construction, clustering, and UMAP visualization.

Clustering was performed iteratively across multiple resolution parameters to assess cluster stability and biological interpretability. Cluster labels from the sketched dataset were subsequently projected onto the full dataset using Seurat’s ProjectData workflow, using the first 30 principal components to transfer annotations and generate full-dataset visualizations while retaining the computational efficiency of sketch-based analysis. The final clustering resolution was selected to maximize identification of biologically distinct cellular populations while minimizing over-fragmentation.

Initial clustering did not fully resolve expected T cell and B cell subpopulations. Therefore, T cell and B cell compartments were independently reanalyzed using an additional sketch-based workflow with iterative sub-clustering and annotation. Refined subtype annotations were subsequently transferred back to the full dataset using anchor-based mapping. Cells in clusters containing indistinguishable CD4 and CD8 markers were assigned to one population or the other based on scoring against custom gene expression programs. Labeling was collapsed into broad cell groups and further refined by overall expression. All downstream analyses were performed using the resulting integrated and annotated dataset. Uniform Manifold Approximation and Projection (UMAP) was used for visualization of cellular relationships.

For visualization of gene expression across cell types, pseudobulk count matrices (sample-by-cell-type) were generated using Seurat’s AggregateExpression function. To correct for differences in library size and RNA composition between samples, pseudobulk counts were normalized using the trimmed mean of M-values (TMM) method implemented in edgeR and transformed to log2 counts per million (logCPM; prior count = 1) using the cpm function (v4.2.2) (Robinson et al., 2010). Heatmaps were generated using ggplot2 and display gene expression across donor-cell-type pseudobulk profiles.

Differentially expressed genes (DEGs) between control and patient cell subsets were identified using a pseudobulk approach implemented in DESeq2 (v1.44.0) (Love et al., 2014). For each annotated cell subset, transcript counts were summed across cells to generate pseudobulk count matrices. Counts from technical replicate sections were first aggregated within each ROI. For control samples, a single ROI was analyzed per subject, yielding one pseudobulk profile each for C1 and C2. To avoid pseudoreplication for the GPR174-deficient patient, counts from three independently sampled ROIs were aggregated to generate a single patient-level pseudobulk profile. The final differential expression analysis therefore compared two control subject-level pseudobulk profiles (C1 and C2) with one patient-level pseudobulk profile (P2). Genes expressed in more than 10% of cells in at least one condition were retained for analysis. Differential expression testing was performed using the DESeq2 Wald test. Given the limited number of biological replicates, DESeq2-derived adjusted p-values were interpreted as exploratory measures of statistical support rather than definitive estimates of population-level significance.

Gene set enrichment analysis (GSEA) was performed using the fgsea R package (v1.35.6) (Korotkevich et al., 2021). A ranked gene list was generated using a hybrid metric defined as the product of the DESeq2 log2 fold change and −log10(adjusted p-value). Replication of the analysis using only effect sizes (log2 fold changes) for ranking generated similar results. Enrichment analysis was conducted against the MSigDB Hallmark gene sets obtained via the msigdbr package (v26.1.0) (Dolgalev, 2026).

Normalized enrichment scores were calculated using a fast permutation-based approach as implemented in fgsea. Normalized enrichment scores (NES) and Benjamini-Hochberg adjusted p-values were used to assess pathway-level significance. Results were visualized in R using ggplot2 (v4.0.3) (Wickham et al., 2026).

### Mouse studies

C57BL/6 (CD45.2) and CD45.1 (BoyJ) mice originated from Jackson Labs.

GPR174 mice (previously generated at UCSF) were genotyped by flow cytometry for the TdTomato knock-in/knock-out reporter.

Mixed bone marrow chimeras were generated by reconstituting mice irradiated using an X-ray source at 900 cGy with freshly isolated bone marrow of the indicated genotypes. Infection studies were performed after 8-9 weeks of reconstitution.

For LCMV Clone 13 (Cl13) chronic infection studies (Matloubian et al., 1994), CD4 depletion was achieved with two daily doses of 100 µg anti-CD4 antibody (BioXCell, GK1.5) administered intraperitoneally. On the evening of the second day, mice were infected with 3 x 10^6^ pfu of LCMV Cl13. Spleens were collected 21 days later and mechanically dissociated for analysis.

## Author contributions

Conceptualization: YH, KA, SH, NR, JC

Data curation: YH, KA, KRE, HG, MMee, MMiano, FR, AG, MR, IC, GD, JF, MCG, VP, SB, TG, TAB, FM, SH, RB, BA, UB, HT, EB, DAO, ECC

Investigation: YH, KA, HT, JSS, YX

Analysis: YH, KA, SR, KRE, HG, MMee, MR, SB, SH, HT, SO

Visualization: YH, KA, SR, HG

Writing (original draft): YH, KA, NR, JC

Writing (review & editing): YH, KA, HT, JSS, KRE, HG, MMee, MMiano, FR, AG, MR, IC, GD, JF, VP, SB, TG, TAB, FM, SH, RB, BA, UB, HT, EB, SR, DAO, ECC, YX, SO, SH, NR, JC

Supervision: MMiano, SH, NR, JC

## Data Availability

Data from the National Genomic Research Library (NGRL) used in this research are available within the secure Genomics England Research Environment. Access to NGRL data is restricted to adhere to consent requirements and protect participant privacy. Data used in this research include the germline variant calls from standard vcf files and phenotypic descriptions of rare disease and cancer participants from the main programme of the 100,000 Genomes Project. The locations of per participant standard vcf files and human phenotype ontology terms are recorded in Labkey tables available within the Research Environment. Access to NGRL data is provided to approved researchers who are members of the Genomics England Research Network, subject to institutional access agreements and research project approval under participant-led governance. For more information on data access, visit: https://www.genomicsengland.co.uk/research.
Xenium spatial transcriptomics data is being deposited to GEO and an accession number will be provided when available.
Otherwise, sequencing datasets generated and analyzed for this study are not publicly available due to ethical and privacy restrictions, as specified in the informed consent obtained from participants and in accordance with applicable data protection regulations. De-identified data supporting the findings of this study are available from the corresponding author upon reasonable request, subject to approval by the relevant ethics committee. Variants in this cohort will be deposited to ClinVar upon publication.

https://ucsf.box.com/s/g4xt8v22oxy5n2osrwqhg7m8if2943wk

## Acknowledgements

We gratefully acknowledge:
- All patients, families, and healthy donors who participated in this study.
- The participants of the National Genomic Research Library (NGRL), whose contributions made this research possible. Secure access to the NGRL under project ID #19 was provided by Genomics England, which delivers the NGRL in partnership with NHS England, and is wholly owned by the UK Department of Health and Social Care. The NGRL contains participants’ health data collected by the NHS as part of their care, along with samples and data from their participation in research, for which fully informed consent has been obtained. This includes genomic and clinical data provided through the NHS Genomic Medicine Service, as well as data obtained through research studies, including the 100,000 Genomes Project and the Generation Study, both of which are delivered in partnership with the NHS, and from other research cohorts involving external collaborators.
- The UCSF Genomics CoLab and UCSF Center for Advanced Technologies, supported by UCSF PBBR, RRP IMIA, and NIH 1S10OD028511-01 grants for sample preparation and sequencing.
- The Immune Dysregulation Program at the Children’s Hospital of Philadelphia.
- Max Eldabbas, Emileigh Maddox, Tanishk Sinha, and Jiayi Shu of the Human Immunology Core at the Perelman School of Medicine at the University of Pennsylvania for assistance with leukapheresed PBMCs. The HIC is supported in part by NIH P30 AI045008 and P30 CA016520.
- Megan Cooper (WashU) and Xiao Peng (Montefiore) for assistance with patient recruitment, and Scott W. Canna (University of Pennsylvania) for supplies.
- The members of our research groups for thoughtful discussions.

## Funding

This work was supported by the following funding sources:

YH: NIAMS Academic Rheumatology and Clinical Immunology T32 AR079068

MMiano: Ministero della Salute, Italy through 5XMILR19 to M.M. and “Ricerca Corrente” funds to Giannina Gaslini Institute

SB: National Institute for Health and Care Research (NIHR) Manchester Biomedical Research Centre (BRC) (NIHR203308)

DAO: Parker Institute for Cancer Immunotherapy and V Foundation for Cancer Research, via a co-sponsored Parker Bridge Fellow Award

SH: Wellcome (207556_Z_17_Z), Pharming (unrestricted research grant) and MRC (MR/Y013395/1)

NR: National Institutes of Health, National Institute of Allergy and Infectious Disease (AI179680) and the Jeffrey Modell Foundation

JC: NIH R01 AI040098 and the Howard Hughes Medical Institute

## Conflicts of interest

TG: Consulting, advisory work and educational support from BioCryst, CSL Behring, KalVista, Novartis, Octapharma, Otsuka, Pharming, Pharvaris and Takeda.

JGC: Consultant for BeBio Pharma, Lycia Therapeutics, and DrenBio Inc.

## Notes

### Author Declarations

Newcastle and North Tyneside Research Ethics Committee 1 of Newcastle University Translational and Clinical Research Institute gave ethical approval for this work. The Institutional Review Board of Children's Hospital of Philadelphia at University of Pennsylvania gave ethical approval for this work. The Ethical Committee of Liguria Region for IRCCS Instituto Giannina Gaslini gave ethical approval for this work.

