## Supplementary figures and images for "Human GPR174 deficiency drives polyclonal lymphoproliferative disease via defects in T cell function"

### Supplemental Figure 1

**Supplementary Figure 1**

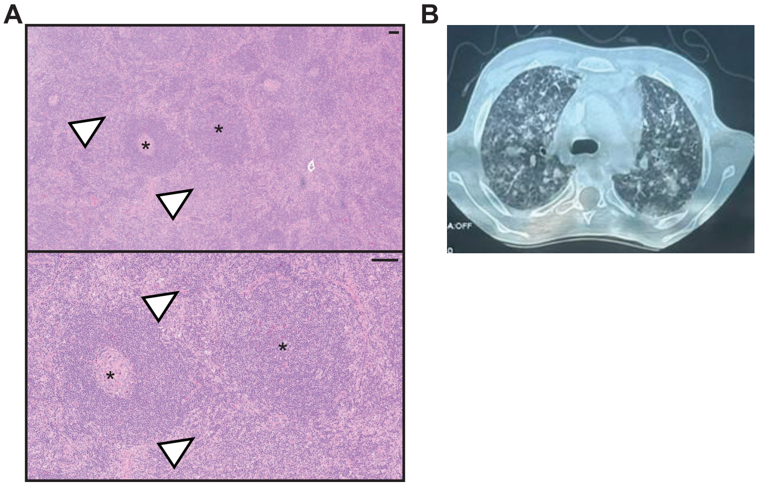

### Supplemental Figure 2

**Supplementary Figure 2**

**A**

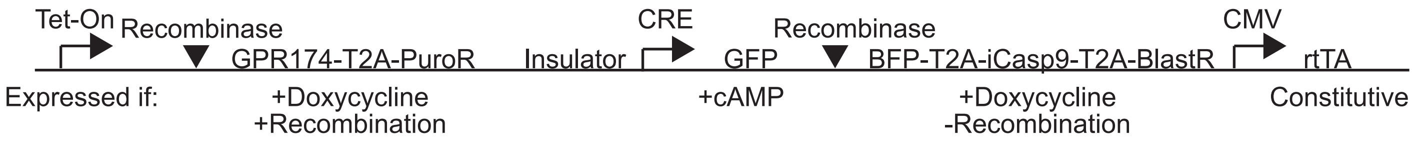

### Supplemental Figure 3

**Supplementary Figure 3**

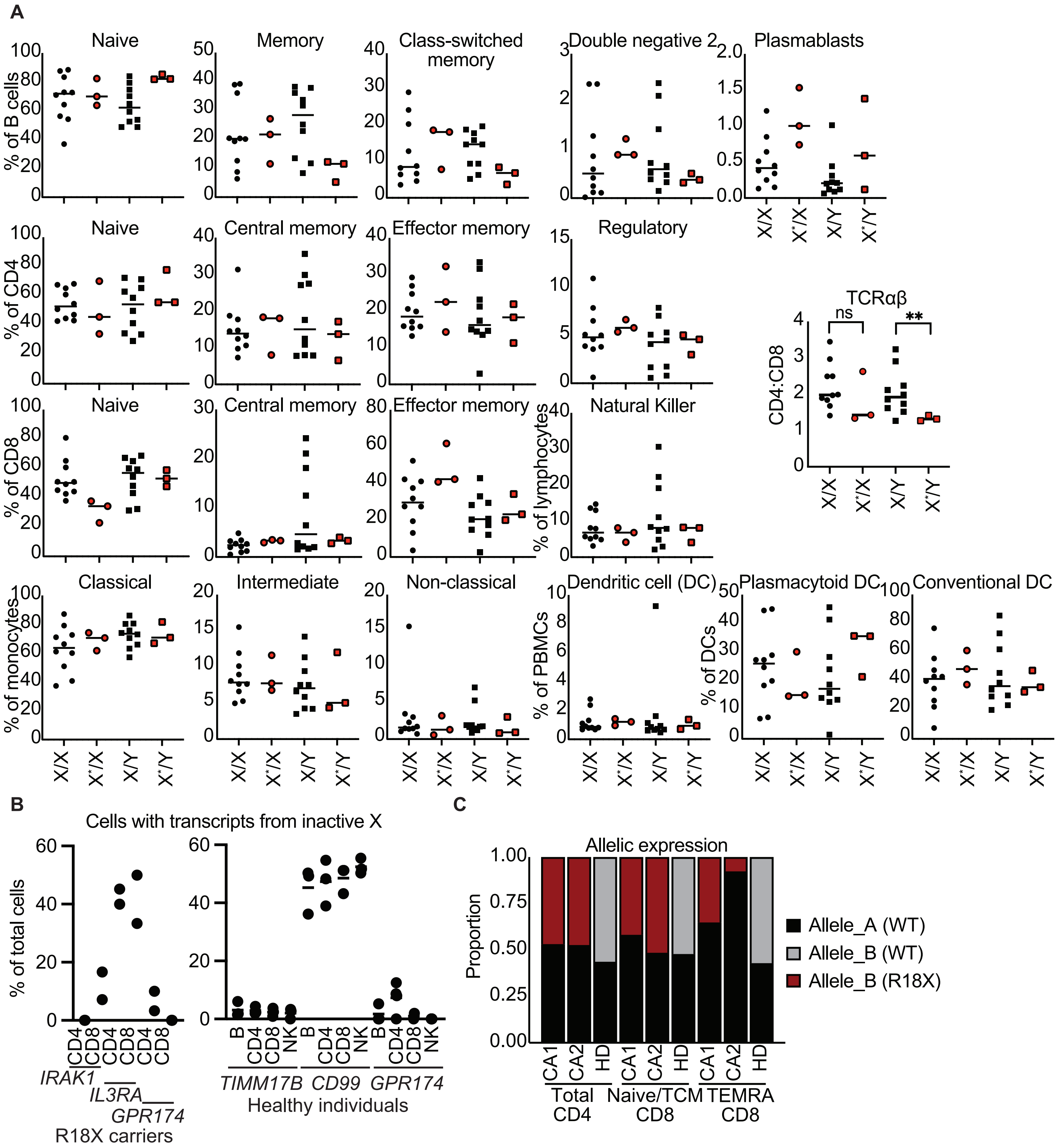

### Supplemental Figure 4

**Supplementary Figure 4**

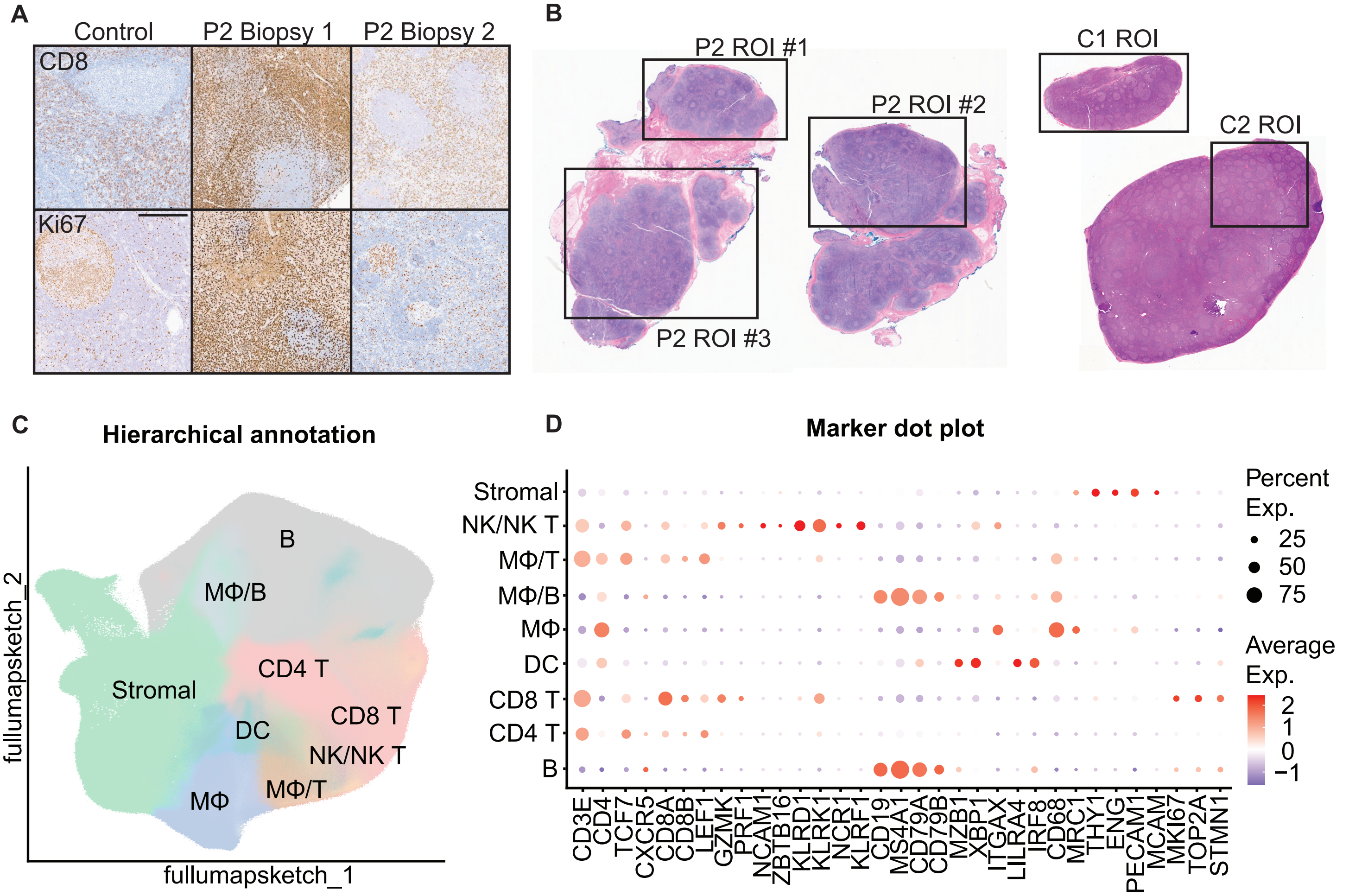

### Supplemental Figure 5

**Supplementary Figure 5**

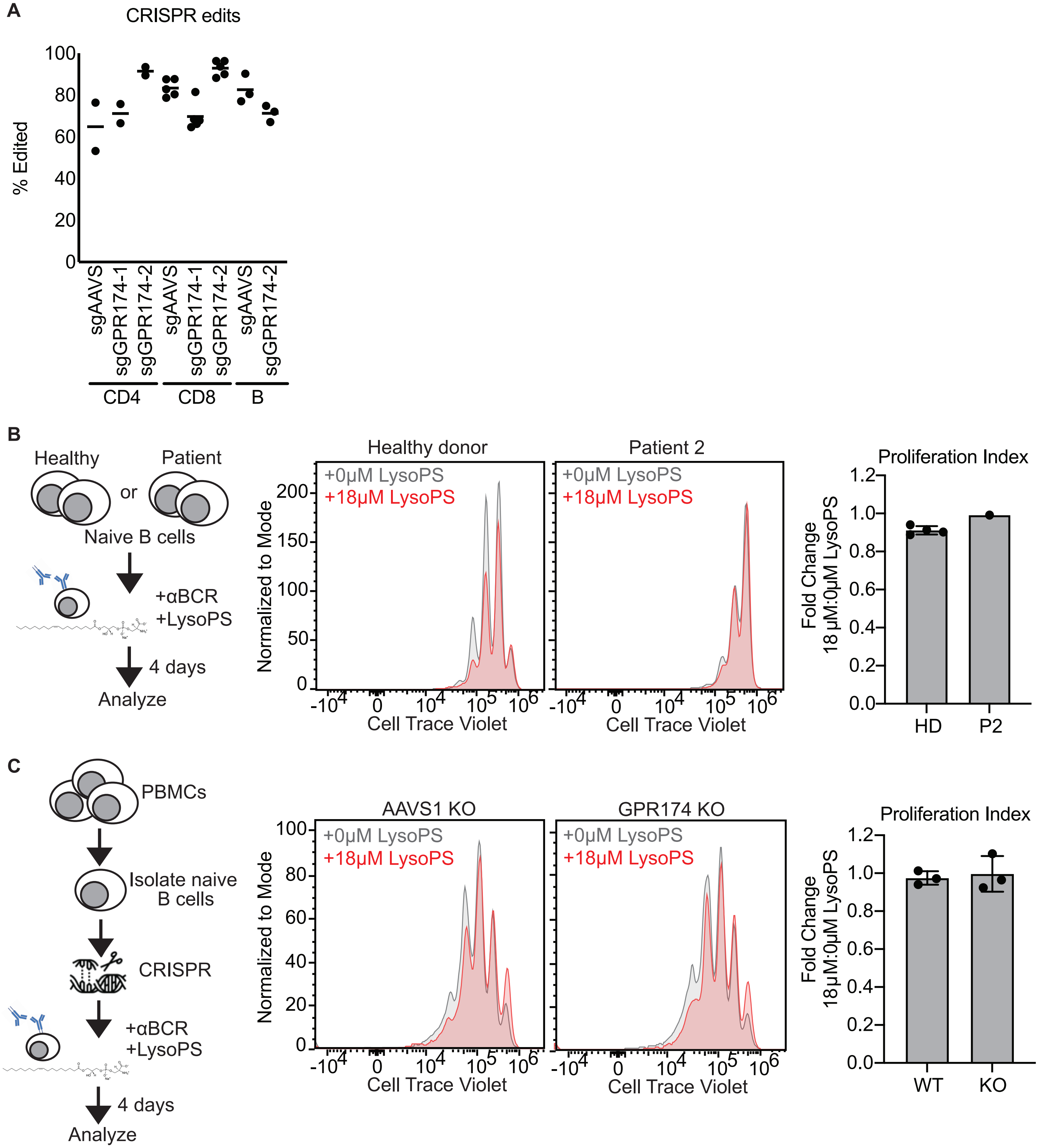
